# A Bayesian latent-class model framework to estimate disease burden of respiratory syncytial virus using imperfect and heterogeneous laboratory diagnostic data

**DOI:** 10.64898/2026.03.24.26349146

**Authors:** Bingbing Cong, Durga Kulkarni, Han Zhang, Caini Wang, Elizabeth Begier, Caihua Liang, Andrew Vyse, Sonal Uppal, Xin Wang, Harish Nair, You Li

## Abstract

**Background:** Accurate estimation of respiratory syncytial virus (RSV) disease burden is challenged by the imperfect testing performance that varies by clinical specimens, diagnostic tests, and timing of specimen collection. Although the use of multiple testing approaches (such as testing multiple clinical specimens or additional diagnostic tests) could increase the RSV detection, there is absence of a modelling framework to fully incorporate the complexity of heterogeneous diagnostic data. In this study, we proposed a novel Bayesian latent class model that accounted for heterogeneous data on the number of RSV tests and variable specimen collection time among individual patients, imperfect testing sensitivity and specificity of different combinations of clinical specimen and diagnostic test (i.e., testing approaches), and RSV seasonality.

**Methods:** Using simulated datasets consisting of four different testing approaches that mimic real-world RSV epidemiologic characteristics in the UK under different sample size and testing practice scenarios, we assessed the model performance in estimating RSV disease burden as the annual RSV positive proportion in lower respiratory tract infection (LRTI) cases across three respiratory seasons (August 2021 to July 2024) in four adult age groups: 18 to 49 years, 50 to 64 years, 65 to 74 years and ≥ 75 years.

**Results:** We demonstrated that model performance increased substantially with increased sample size, achieving over 80% in accuracy at a sample size of 30,000 tests and 95% in accuracy at a sample size of 60,000 tests; by contrast, smaller sample size could lead to severe over-estimation of the RSV disease burden. In comparison with the existing approaches, both the naïve model and the multiplier model systematically under-estimated the RSV disease burden regardless of sample size. The Bayesian model yielded more accurate estimates when the sample size reached 30,000 tests or more; its advantage over the other two models was even more pronounced if the number of testing approaches reduced to 3.

**Conclusion:** The findings above suggest that the proposed Bayesian model provides a robust framework for estimating RSV burden by integrating complex, individual-level testing data when fitting with sufficient input data, offering a critical tool for generating more accurate RSV disease burden estimates to inform national immunisation policies.

**Key messages:**

- We aimed to develop and validate a novel Bayesian latent-class model to accurately estimate RSV disease burden by accounting for variability in test sensitivity and specificity, sampling timing, individual testing patterns, and RSV seasonality that conventional naive and multiplier models fail to fully account for.
- Our Bayesian model demonstrated significantly higher accuracy in estimating RSV burden with increased sample size (over 80% at 30,000 tests, over 95% at 60,000 tests), outperforming naïve and multiplier models that systematically underestimated RSV burden regardless of sample size, with its advantage being more pronounced when the number of testing approaches was reduced.
- This model provides a robust, flexible analytical framework for integrating heterogeneous diagnostic data to generate accurate RSV burden estimates, which is critical for informing RSV immunisation strategies, prioritising high-risk populations, and optimising public health interventions to reduce the impact of RSV-related lower respiratory tract infections.

## Introduction

Respiratory syncytial virus (RSV) is an important cause of lower respiratory tract infection (LRTI) in older adults^1^ and younger adults with underlying comorbidities^2^. However, there is substantial uncertainty regarding the “true” RSV burden in these populations, including both RSV cases detected by existing standard-of-care testing and cases missed that could be identified through additional salvage testing^3^. As three adult RSV vaccines (including RSVPreF3, GSK; RSVPreF, Pfizer and mRNA-1345, Moderna) are now licensed and being introduced into national immunisation programmes globally, primarily focussing on older adults aged ≥75 years^4,5^, adults 50 to 75 years with underlying medical conditions^4,6^, and care home residents^6^, there is an urgent need to accurately estimate RSV disease burden in adults in order to inform the evidence for broadening the scope (including lowering the age thresholds, identifying risk groups etc.) and optimising the immunisation strategy.

However, accurate direct estimation of RSV disease burden in adults is challenging due to several reasons. Firstly, adults are less likely to undergo RSV testing than children in clinical practice, resulting in potential underdiagnosis in the adult population^7^. Secondly, even when RSV testing is conducted, the relatively low viral loads in RSV-infected adults compared to children could lead to false negative results and subsequent under-detection^8^; previous meta-analyses reported that compared to a single RT-PCR test of nasal or nasopharyngeal swab, addition of another specimen paired serology would increase the detection of RSV by 42% in adults whereas the increase in children was limited^9,10^. The third challenge is the rapid decline of viral loads following the first five days after symptom onset, leading to a lower sensitivity when specimen collection was not conducted in time^11^. Moreover, studies that collected four specimens, nasopharyngeal swab, saliva, sputum and serum (for serology) showed that each of the specimen types added unique RSV detections^12,13^; compared to nasopharyngeal swab alone, RSV detection was 112% higher when all four specimens were combined^12^. These challenges inherent to limited laboratory testing highlight the need for appropriate statistical adjustment for a better understanding of the RSV disease burden in adults.

To adjust for the potential under-estimation of RSV disease burden, existing studies have applied multipliers at the population level to upscale the RSV incidence. For example, McLaughlin and colleagues applied a multiplier of 1.5 to account for the under-detection when using nasopharyngeal specimen alone compare to adding sputum or serology^14^. Likewise, in a systematic review and meta-analysis, Li and colleagues developed a two-step approach to adjust for the use of clinical specimens and diagnostic tests in different studies, resulting an average multiplier of 2.2^15^. The US CDC also developed a multiplier method to adjust for case under-ascertainment due to testing practices over time and test sensitivity^16^. However, the use of multipliers in these studies over-simplified the adjustment by omitting a number of potentially important factors. First, all adjustments were conducted at the study or population level rather than at the individual level, thus not accounting for the potential heterogeneity in specimen collection time and testing approaches among individuals. Second, existing multi-specimen studies assumed that all RSV cases could have been identified by the prespecified gold standard such as the combination of four specimens (e.g., nasopharyngeal swab, saliva, sputum and serum) while it was possible that adding an additional further specimen could yield additional RSV detection (e.g., bronchoalveolar lavage), leading to underestimation of the true disease burden. Third, although it was well acknowledged that the specificity of RSV diagnostic tests was close to 100%, use of a single upscaling factor without adjusting for false positive means that the existing studies theoretically over-estimated the RSV disease burden, particularly when RSV circulation was low (leading to a lower positive predictive value of RSV tests).

To overcome these challenges, we proposed a novel Bayesian latent-class model framework to explicitly account for heterogeneous data on the number of RSV tests and specimen collection time at the individual level while adjusting for imperfect sensitivity and specificity of different clinical specimens and diagnostic tests as well as RSV seasonality. By simulating datasets under different sample size and testing practice scenarios, we aimed to assess the accuracy of the model and identify scenarios where the model could be applied to yield robust estimates of RSV disease burden.

## Materials and methods

### Proposed model

The proposed model included a latent class analysis indicating RSV infection status of an individual *i* who met the criteria for RSV testing (e.g., with LRTI). Here, we assumed that the probability of RSV infection was determined by the internal factor of age *a* (as four age groups: 18 to 49 years, 50 to 64 years, 65 to 74 years and ≥ 75 years) and the external factor of RSV circulating strength denoted as calendar month *m* for a specific year to account for RSV seasonality; *θ*_*m,a*_ represented the proportion of RSV infection among LRTI cases in a given age group *a* and month *m* among the study population and was the key parameter for estimating RSV disease burden. The corresponding formula is given below:

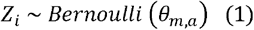

Individual *i* might provide up to four different specimens for RSV diagnostic tests (referred to as testing approach *j*) with likely time lag in specimen collection resulting in time-varying sensitivity; here, we used the term peak sensitivity for referring the maximum sensitivity of a testing approach *j*, denoted as *Sens* _*j*_, and used the term relative sensitivity for referring to the change of sensitivity over specimen collection time denoted as *W*_*t*_; we assumed that specificity *Spec*_*j*_ remained constant. Then the RSV positivity result *Y*_*ij*_ by a specific testing approach *j* in an individual *i* could be denoted as:

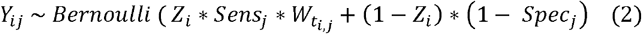

Here, peak sensitivity *Sens*_*j*_ is an unknown parameter for inference; for the specimen collection time, we prespecified four time slots, [0, 2], (2, 5], (5, 6] and (6, 14] days after presentation to the health-care, with the corresponding relative sensitivity *W*_*t*_ assumed to be 1, 0.8, 0.6 and 0.5, respectively; specificity *Spec* _*j*_ for each testing approach was assumed to be known and fixed at 0.998, based on the reported high specificity of existing RSV molecular testing approaches^9^.

Therefore, the annual disease burden for a season *s* as the number of RSV-associated LRTI cases, could be estimated by totalling up the number of RSV-associated LRTI cases across the 12 months, as follows –

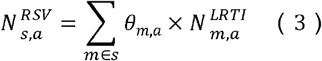

Lastly, the annual proportion of RSV in LRTI cases for that season could be calculated as:

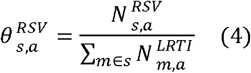

### Simulation

To enhance the real-world applicability of the model framework, we selected to simulate RSV testing datasets across three respiratory seasons (August 1, 2021 to July 31, 2024) using adults in the UK as the population under study, and derived parameters from real-world evidence in the UK for the contemporaneous period. We defined each season as 12 calendar months from August 1 to July 31 of the following year given that RSV epidemics usually occurred during the autumn and winter months in the UK^17^. Detailed parameters and specifications for the simulated datasets are summarised in **Table 1**. Briefly, we referred to the AvonCAP multi-specimen study^18^ that applied four RSV testing approaches among adults hospitalised due to LRTI for parameters regarding distribution of number of RSV testing approaches across LRTI cases; we referred to surveillance data in the UK^19^ for the distribution of LRTI cases by calendar month and a published study^20^ on age-specific LRTI hospitalisation rate for the distribution of LRTI cases by age group.

**Table 1.** Key parameters applied to the simulation.

| Parameters | Specification |
| --- | --- |
| **Parameters set as fixed values** |  |
| Data period | Three respiratory seasons from August 1, 2021 to July 31, 2024 (i.e., August 1, 2021 to July 2022, August 1, 2022 to July 2023, August 1, 2023 to July 2024) |
| Age groups | 18 to 49 years, 50 to 64 years, 65 to 74 years and ≥ 75 years |
| Distribution of LRTI patients by age group | 14%, 15%, 23%, and 48% for 18 to 49 years, 50 to 64 years, 65 to 74 years and ≥ 75 years, respectively, based on the differences between LRTI hospitalisation rate<sup>20</sup> and population size among age groups<sup>28</sup>. |
| Distribution of LRTI patients by calendar month | Based on LRTI seasonality data from surveillance in the UK<sup>19</sup>. (details in Figure S3). |
| Distribution of number of tests per patient | 1 test: 42%; 2 tests: 28%; 3 tests: 18%; 4 tests: 12% (based on distribution in the AvonCAP multi-specimen study<sup>18</sup>). |
| Distribution of specimen collection time (days since admission) | [0, 2]: 37%; (2, 5]: 30%; (5, 6]: 22%; (6, 14]: 11%. |
| Relative sensitivity of tests at different time of specimen collection (days since admission) | [0, 2] days: 100%; (2, 5] days: 80%; (5, 6] days: 60%; (6, 14], 50%<sup>11</sup>. |
| Specificity of tests | 0.998<sup>9</sup> |
| **Parameters varied by scenario** |  |
| Sample size: LRTI patients | 2,500; 5,000; 7,500; 10,000; 15,000; 20,000; 30,000 |
| Distribution of tests by testing approach | **Main scenario:** proportional to the sensitivity — 31%, 27%, 23%, and 19% for Tests 1–4;<br>**Alternative scenario 1:** equal distribution (i.e., 25% for all);<br>**Alternative scenario 2:** inversely proportionally to the sensitivity — 19%, 23%, 27%, and 31% for Tests 1–4. |
| Distribution of tests by calendar month | **Main scenario:** same to the monthly proportion of patients;<br>**Alternative scenario 3:** equal distribution regardless of number of patients;<br>**Alternative scenario 4:** disproportionately higher proportion during high season.<br>(details in Figure S4) |
| **Parameters for inference (set as fixed values in the simulation)** |  |
| Testing approach (true sensitivity) | Test 1 (80%), Test 2 (70%), Test 3 (60%), Test 4 (50%) |
| True proportion of RSV infection in LRTI patients by age group and calendar month | Based on RSV seasonality data from surveillance in the UK<sup>29</sup> and a previous study reporting RSV positive proportion in different age groups<sup>18</sup>.<br>(details in **Text S1** and **Figure S1**) |

RSV: respiratory syncytial virus; LRTI: lower respiratory tract infection.

The true values of parameters for inference for generating simulated datasets included the peak sensitivity of the four tests, which were hypothetically set as 80%, 70%, 60%, and 50%, representing the plausible range of sensitivity, and the age- and calendar-month specific RSV positive proportions, which were calibrated to real-world evidence on RSV activity during the simulation time period in the UK, ranging from 0 to 39.7% (details in **Text S1** and **Figure S1, appendix**).

To account for the potential variability around testing practices, specifically how the number of tests across different testing approaches and months of a year were determined, we considered a total of five testing practice scenarios, including one main scenario that assumed the number of tests was proportional to the sensitivity of the test (i.e., tests with greater sensitivity were used more frequently) and proportional to the number of LRTI patients per month (i.e., individuals had the same chance of receiving the test regardless of calendar month), and four alternative scenarios (detailed in **Table 1** and **Figure S2**). For each testing practice scenario, we considered a total of seven sample size scenarios, defined as the number of LRTI patients receiving at least one RSV test: 2,500, 5,000, 7,500, 10,000, 15,000, 20,000 and 30,000; for all sample size scenarios, we assumed that the total number of RSV tests was twice the number of LRTI patients.

For each combination of testing practice scenarios and sample size scenarios, we simulated four datasets; the total number of simulated datasets was 5 testing practice scenarios * 7 sample size scenarios * 4 datasets per scenario = 140.

### Bayesian inference

For all simulated datasets, we implemented Bayesian inference through the Stan software. We used non-informative beta distribution priors for inferring unknown parameters, including the age- and calendar-month specific RSV positive proportion, and the peak sensitivity of testing approaches, as follows –

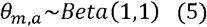

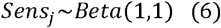

Model parameters were estimated by using the No-U-Turn sampler of Hamiltonian Monte Carlo implemented through the rstan package^21^ in R 4.2.1. For each scenario, we ran the same model with 6,000 iterations and a burn-in period of 3,000 iterations, in each of 4 chains. Convergence was assessed using Rhat estimates (Rhat < 1.05 indicating convergence) and visually through trace plots.

### Model assessment

We assessed the model calibration performance using the slope of the calibration curve between the pre-specified true values and Bayesian model estimates, fitted with linear regression models with no intercepts; perfect calibration is indicated by a slope of 1, whereas a slope above 1 indicates over-estimation of parameters and a slope below 1 indicates under-estimation. We also assessed whether the estimated 95% credible intervals (CrIs) contained the true value for each simulated dataset, and compared the proportion of estimates that contained the true value across different scenarios; when the estimated CrIs did not contain the true value, we further defined under-estimation [over-estimation] as the estimated CrIs being below [above] the true value. The assessment above was conducted for each parameter for inference, *Sens* _*j*_ and *θ*_*m,a*_ as well as the annual proportion of RSV, 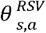. The full workflow is summarised in **Figure 1**.

**Figure 1.**
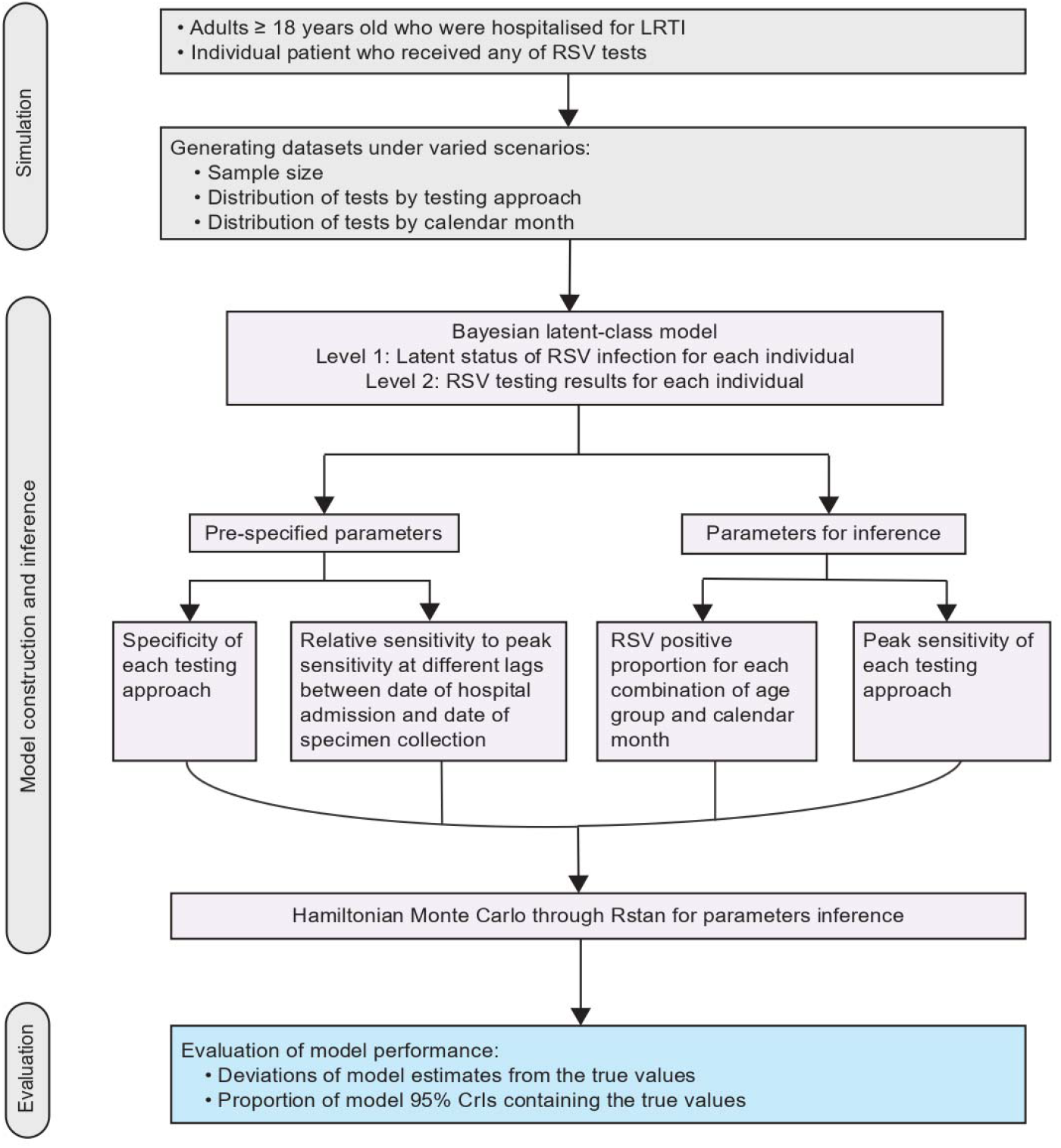
Schematic figure of methods. RSV: respiratory syncytial virus; LRTI: lower respiratory tract infection; 95% CrIs: 95% credible intervals.

### Comparison with existing approaches

We further compared the Bayesian model estimates with those obtained by the existing approaches, namely the naïve approach and the multiplier approach.

For the naïve approach, no adjustments were made according to the specific testing status of an individual; an individual who tested positive for RSV from any of the testing approaches was regarded as positive and an individual who tested negative for RSV from all testing approaches available was regarded as negative. As in the Bayesian model, we first estimated the RSV positive proportion for each calendar month per age group, *θ*_*m,a*_ and then followed formula (3) and (4) to estimate the age-specific annual RSV positive proportion.

For the multiplier approach, we regarded individuals who received all the four testing approaches and tested positive for RSV from any of these approaches as a “practical” gold standard (differing from the “true” gold standard as the four approaches might have missed RSV infected cases), and estimated the practical sensitivity of each possible combination of testing approaches by the ratio of RSV positive proportion among patients who received that combination of tests and those who received all the tests (**Table S1, appendix**). The reciprocal of the practical sensitivity estimates was then used as multipliers to upscale the RSV positive proportion to yield the RSV positive proportion for each calendar month per age group (*θ*_*m,a*_), and the age-specific annual RSV positive proportion as done in the naive approach.

As an exploratory analysis, we reduced the number of testing approaches to 3 (retaining testing approaches 1, 2, and 3) and repeated the comparison above under the same sample size scenarios.

## Results

### Description of simulated datasets

The true RSV positive proportion in the simulated datasets varied from 4.8% to 6.5% in adults aged 18 to 49 years; 4.4% to 10.1% in adults aged 50 to 64 years; 4.1% to 9.9% in adults aged 65 to 74 years; and 3.4% to 14.8% in adults aged ≥ 75 years, across the three seasons. Overall, the observed nominal positive proportions, defined as the presence of any positive test results, were generally lower than the corresponding true positive proportions (**Table 2**).

**Table 2.** Nominal RSV positive proportion in LRTI cases across sample size scenarios by age group and season from the simulated datasets.

|  | 2021 to 2022<br>season | 2022 to 2023<br>season | 2023 to 2024<br>season |
| --- | --- | --- | --- |
| **18 to 49 years** |  |  |  |
| True positive proportion (%) | 4.8 | 6.2 | 6.5 |
| Range of nominal positive<br>proportion (%) by sample size |  |  |  |
| 2,500 | 1.2–4.9 | 3.1–8.6 | 3.1–5.4 |
| 5,000 | 1.8–4.2 | 2.0–7.1 | 2.3–4.3 |
| 7,500 | 2.8–3.6 | 4.0–6.3 | 4.2–6.0 |
| 10,000 | 3.0–3.9 | 2.8–4.3 | 2.9–5.1 |
| 15,000 | 3.0–4.2 | 2.9–4.7 | 2.3–3.6 |
| 20,000 | 2.2–2.8 | 3.5–5.3 | 2.7–4.0 |
| 30,000 | 2.9–4.0 | 3.4–4.3 | 2.9–4.0 |
| **50 to 64 years** |  |  |  |
| True positive proportion (%) | 4.4 | 10.1 | 8.4 |
| Range of nominal positive<br>proportion (%) by sample size |  |  |  |
| 2,500 | 2.2–5.4 | 5.7–7.9 | 6.4–7.8 |
| 5,000 | 1.6–6.5 | 6.4–8.2 | 4.6–7.7 |
| 7,500 | 1.1–4.0 | 5.7–9.8 | 4.5–6.1 |
| 10,000 | 1.6–3.0 | 5.7–6.6 | 6.0–6.7 |
| 15,000 | 3.2–4.7 | 6.5–6.9 | 5.1–6.1 |
| 20,000 | 2.0–4.2 | 5.0–7.5 | 5.0–6.8 |
| 30,000 | 2.8–3.1 | 6.6–7.0 | 4.8–6.3 |
| **65 to 74 years** |  |  |  |
| True positive proportion (%) | 4.1 | 9.9 | 7.8 |
| Range of nominal positive<br>proportion (%) by sample size |  |  |  |
| 2,500 | 2.1–2.8 | 3.2–10.1 | 3.7–5.5 |
| 5,000 | 2.4–4.2 | 5.8–8.1 | 3.4–5.9 |
| 7,500 | 2.1–2.8 | 5.2–7.2 | 4.2–5.9 |
| 10,000 | 2.1–3.2 | 5.8–7.6 | 4.8–5.6 |
| 15,000 | 2.0–2.8 | 5.6–7.4 | 4.8–5.5 |
| 20,000 | 2.1–2.9 | 5.5–6.6 | 4.9–6.5 |
| 30,000 | 2.2–3.0 | 5.8–7.3 | 4.1–5.9 |
| **≥ 75 years** |  |  |  |
| True positive proportion (%) | 3.4 | 14.8 | 10.5 |
| Range of nominal positive |  |  |  |
| proportion (%) by sample size |  |  |  |
| 2,500 | 2.0–2.7 | 8.6–11.2 | 5.7–6.8 |
| 5,000 | 2.4–3.4 | 10.3–11.5 | 5.4–7.8 |
| 7,500 | 2.0–2.8 | 9.4–10.8 | 7.3–8.8 |
| 10,000 | 2.5–3.0 | 9.6–10.7 | 6.3–8.0 |
| 15,000 | 2.0–2.9 | 9.1–11.3 | 6.5–7.6 |
| 20,000 | 2.1–2.9 | 9.3–11.3 | 7.1–7.5 |
| 30,000 | 2.2–2.8 | 9.8–10.1 | 7.2–7.6 |

RSV: respiratory syncytial virus; LRTI: lower respiratory tract infection. Each respiratory season begins on 1<sup>st</sup> of August and ends on 31<sup>st</sup> of July. Nominal positive proportion was defined as RSV infection by the presence of positive test results by any of the four tests. Four simulated datasets were generated for each sample size scenario.

### Model performance

Under the main testing practice scenario for the Bayesian model, the calibration curve revealed a marked improvement in model calibration performance with increasing sample size. The modelled peak sensitivity for four tests closely aligned with the true values when sample size reached 20,000 tests, with a calibration slope ranging from 0.81 to 0.97 for all testing approaches. In contrast, insufficient sample size could lead to severely underestimated sensitivity (**Figure 2**).

**Figure 2.**
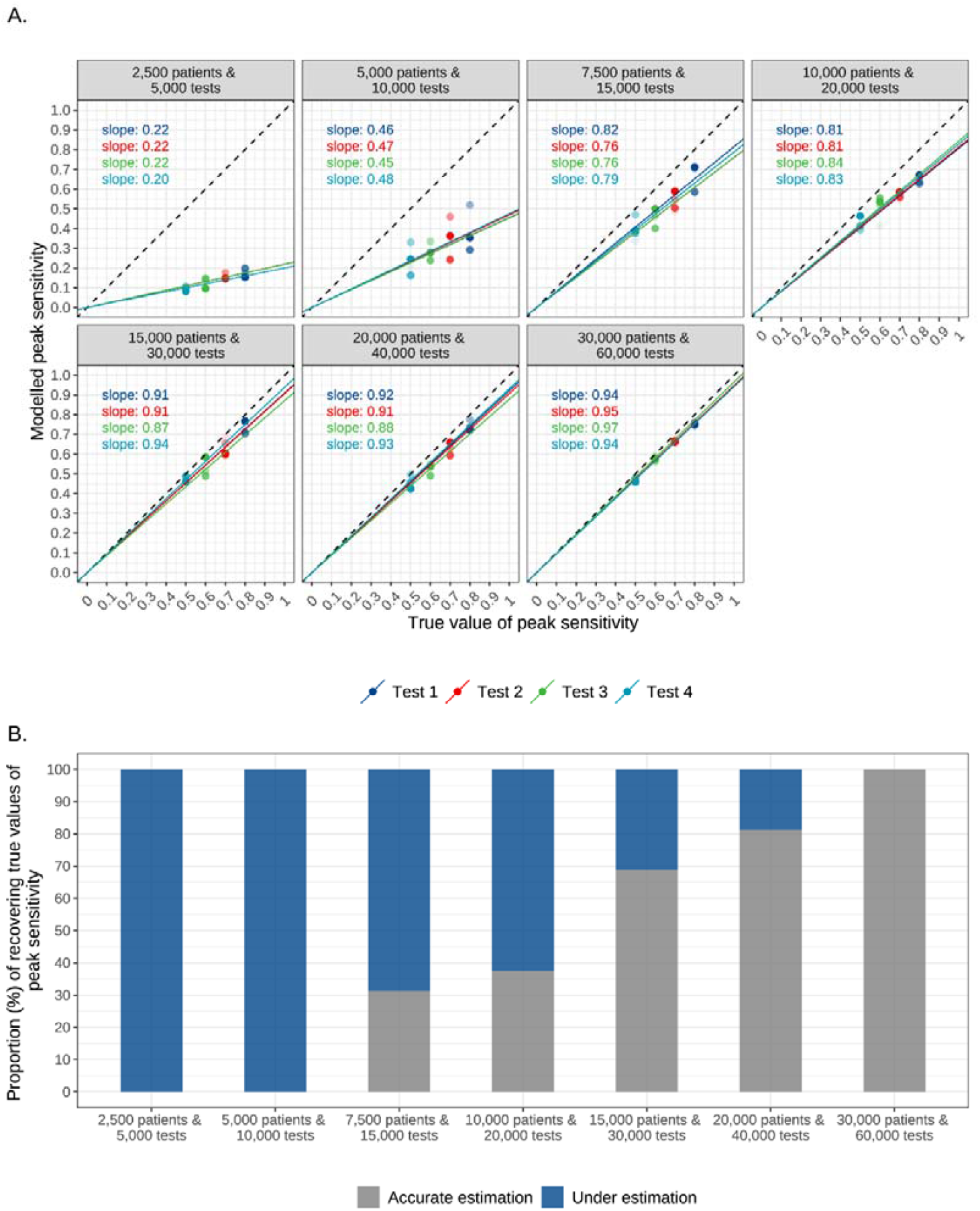
Comparison of model estimates and true values of peak sensitivity of tests under varied sample size scenarios. Four simulated datasets were generated for each sample size scenario. Accurate estimation is defined as the 95% credible intervals containing the true values; under [over] estimation denotes the upper [lower] limit of the 95% credible intervals being lower [higher] than the true value.

Accordingly, the age- and calendar-month specific RSV positive proportion also demonstrated better calibration as sample size increased, with a calibration slope decreasing from 3.60 to 1.07. Overall, over-estimation was observed more frequently than under-estimation. When the sample size was 20,000 tests, the proportion of over- or under-estimation fell below 5%, which further decreased as the sample size increased (**Figure 3**).

**Figure 3.**
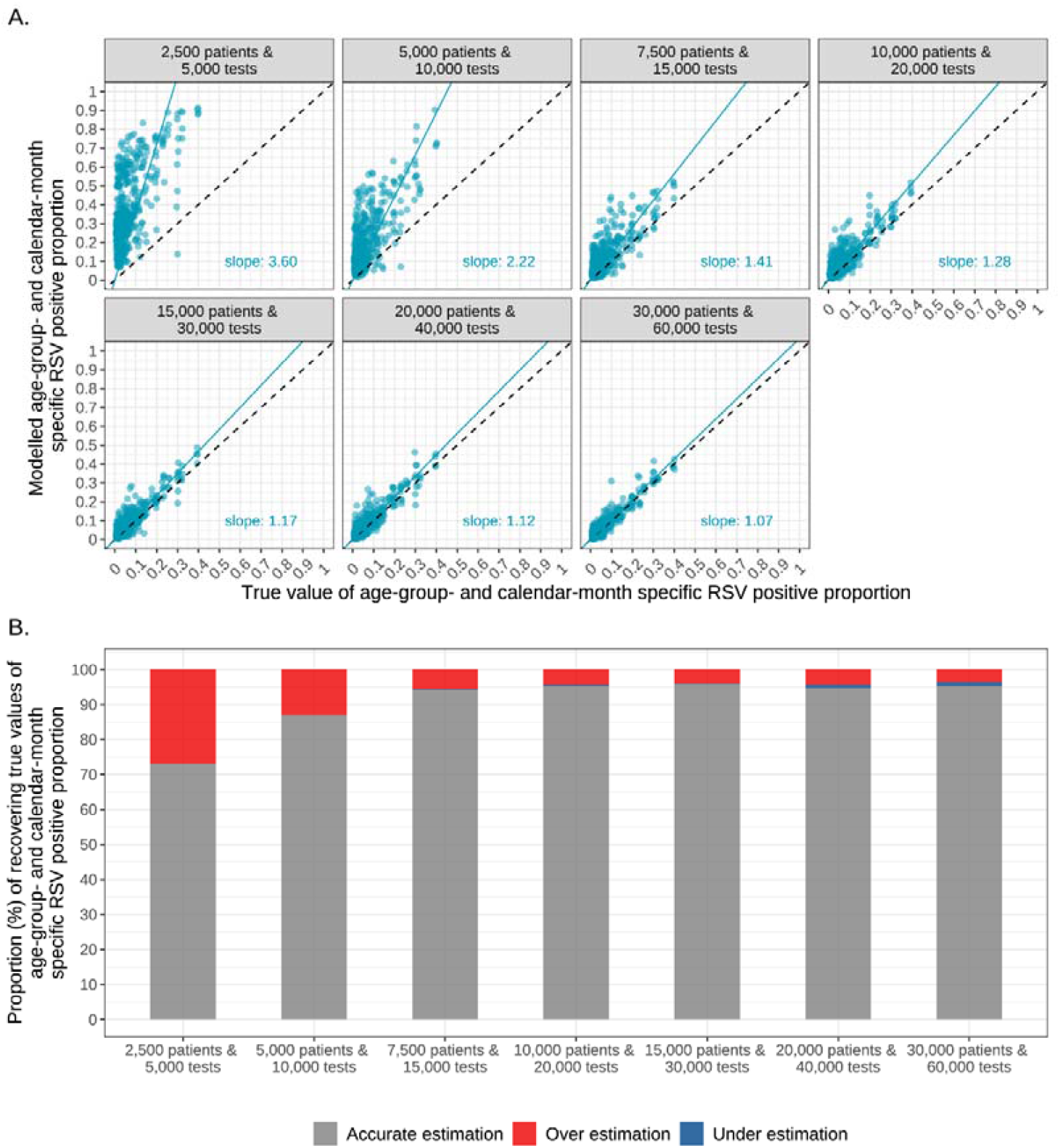
Comparison of model estimates and true values of age- and calendar-month specific RSV positive proportion under varied sample size scenarios. Four simulated datasets were generated for each sample size scenario. Accurate estimation is defined as the 95% credible intervals containing the true values; under [over] estimation denotes the upper [lower] limit of the 95% credible intervals being lower [higher] than the true value.

Similar patterns were observed when aggregating the RSV positive proportion annually by age group. When the sample size increased to 30,000 tests, the proportion of accurate estimation reached 80%; the proportion of accurate estimation further increased to 95% at a sample size of 60,000 tests (**Figure 4**).

**Figure 4.**
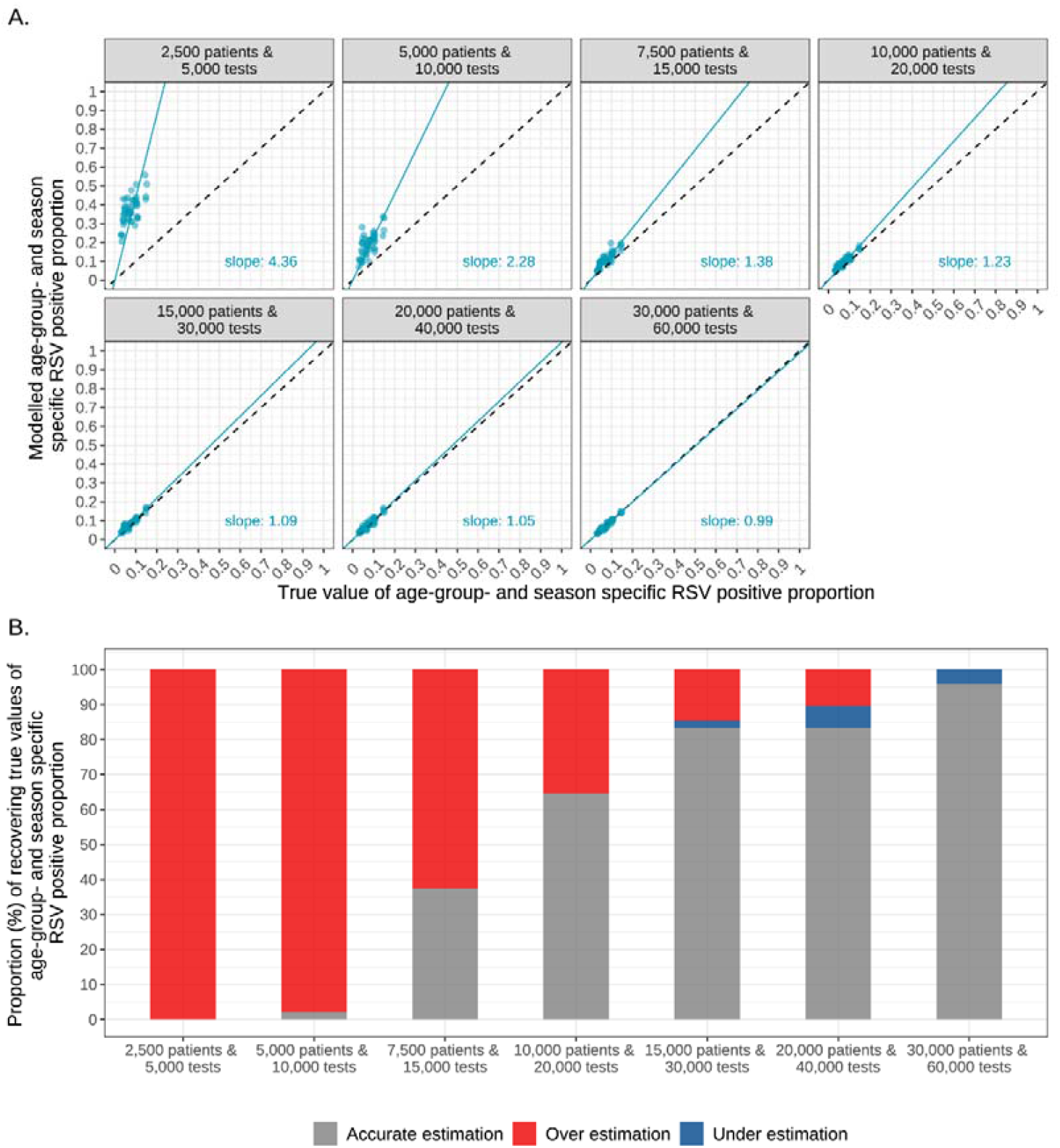
Comparison of model estimates and true values of age- and season-specific RSV positive proportion under varied sample size scenarios. Four simulated datasets were generated for each sample size scenario. Accurate estimation is defined as the 95% credible intervals containing the true values; under [over] estimation denotes the upper [lower] limit of the 95% credible intervals being lower [higher] than the true value.

Additional analyses that used alternative testing practice scenarios yielded findings consistent with the main scenario (**Figures S6–11, appendix**). In the exploratory analysis that retained 3 testing approaches, we found similar patterns for the age- and season-specific RSV positive proportion (**Figure S15, appendix**).

### Comparison with existing approaches

Across the four age groups, the naive model had consistent under-estimation across all sample sizes, with the ratios of model estimates to the true values ranging from 0.38 to 0.88. Although the multiplier model showed moderate improvement in estimating annual RSV positive proportion, under-estimation persisted across all sample size scenarios. Compared to the existing two approaches, the Bayesian model exhibited severe over-estimation at small sample sizes (≤ 15,000 tests); as sample size increased to 30,000 tests, the model demonstrated superior accuracy compared to the other two approaches (**Figure 5**). Stratified analyses by seasons revealed a consistent finding with the three-season aggregated results (**Figures S12–14, appendix**). When reducing the number of testing approaches to 3, the advantage of the Bayesian model was even more pronounced when the sample size was 30,000 tests or above (**Figure S16, appendix**).

**Figure 5.**
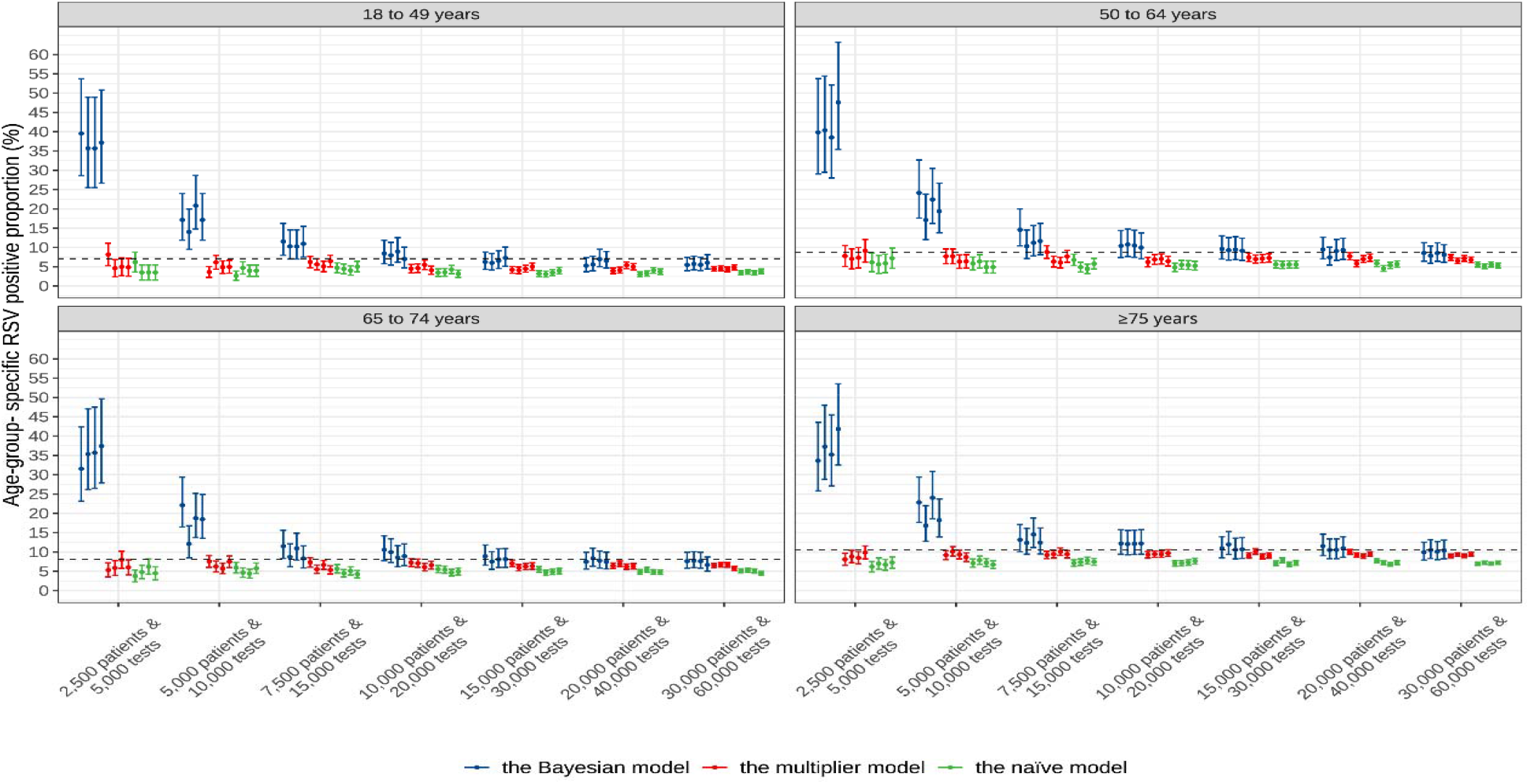
Comparison of age-specific annual RSV positive proportion from August 1, 2021 to July 31, 2024 among the Bayesian model, naïve model, and multiplier model under varied sample size scenarios. Four simulated datasets were generated for each sample size scenario. Dotted lines represent true values of RSV positive proportions. Dots indicate modelled point estimates and corresponding error bars indicate corresponding 95% credible intervals or confidence intervals.

## Discussion

We developed a novel Bayesian latent-class model to systematically address critical challenges in estimating RSV disease burden from laboratory testing data, including variability in clinical specimens, diagnostic tests, specimen collection time, and individual testing patterns. Through a series of scenario analyses that closely matched the real-world RSV epidemiologic characteristics of real-world data, we found that the Bayesian model performance improved with an increase in sample size. When the sample size reached 30,000 tests or more, the Bayesian model demonstrated high accuracy in estimating annual RSV positive proportion in LRTI cases across all age groups and seasons, superior to the existing naïve model and multiplier model, both of which systematically under-estimated the RSV disease burden irrespective of sample size. Furthermore, the proposed Bayesian model suggested improved RSV disease burden estimation with large and heterogeneous RSV diagnostic data.

As expected, the conventional naïve approach consistently under-estimated RSV positive proportions across all sample sizes, reflecting the well-documented limitation that it relied solely on nominal test results without accounting for imperfect test sensitivity or heterogeneous tests among individuals. While the multiplier approach showed moderate improvement by incorporating individual-level adjustment for imperfect test sensitivity, it failed to eliminate under-estimation bias as it relied on a “practical” gold standard (i.e., individuals with positive tests out of a finite number of tests), which was not 100% sensitive. Furthermore, the multiplier approach could not account for the varied specimen collection time of individuals, leading to more uncertainty when variations in specimen collection time were substantial among the individual patients.

Compared to the two existing approaches, the proposed Bayesian model could simultaneously effectively account for imperfect sensitivity and specificity of different testing approaches, and varied testing patterns in terms of the number of testing approaches and specimen collection time among individual patients. However, the full adjustment of all factors above was limited by sample size. We showed that the estimation could be severely biased upwards when the sample size was small (≤15,000 tests across three seasons); specifically, the model could not reliably attribute a low number of detected RSV cases to imperfect test sensitivity versus a truly low level of RSV prevalence in the case of sparse data. As the sample size increased to 30,000 tests or more, the Bayesian model could effectively separate these factors, mitigate the bias and demonstrate its robustness for real-world data with complex measurement errors^22,23^.

This assessment above provides a critical practical guideline for the design of studies applying multiple testing approaches to understand disease burden of a pathogen such as RSV with imperfect testing performance: a minimum of 15,000 tests (corresponding to 7,500 patients) should be considered and the sample size is ideally above 30,000 tests (corresponding to 15,000 patients) for yielding a robust estimation of the disease burden. When necessary, additional simulation and evaluation exercises as in this study might be used to estimate the appropriate sample size when the key parameters or assumptions such as the prevalence of RSV and the test sensitivity vary substantially from those used in this study. For the less ideal scenario of insufficient sample size in a study applying multiple testing approaches, the multiplier approach might be used as a conservative estimation of the disease burden provided that all innate limitations of the approach are well acknowledged.

A primary strength of this study lies in the comprehensiveness of the model design, complementing and extending existing research on RSV disease burden estimation methods. Through explicit assessment of the model performance and comparison with conventional approaches (naïve and multiplier approaches), this study demonstrated the issue of under-estimation in the conventional approaches and the advantages of the Bayesian model under specific conditions. At the time of drafting our manuscript, a Bayesian latent-class analysis model framework was published that accounted for imperfect test sensitivity and specificity of multiple testing approaches to infer true prevalence^18^; however, the authors did not account for the variations of test sensitivity by specimen collection time, as was done in our study.

Our study has several limitations. First, by assuming a fixed and near-perfect specificity, our model prioritised statistical power for the estimation of sensitivity across different testing approaches. While high specificity is well accepted for molecular assays, cross-contamination or non-specific amplification in practice can lead to false positives^24^. Second, parameters regarding the relative sensitivity according to the time of specimen were pre-specified based on previous literature^11^; although such pre-specification reflects the general understanding of declining viral titers, the true values of these factors may vary by individual characteristics, treatment and co-infection status. An alternative approach for improvement was to model relative sensitivity as a function of time since symptom onset based on longitudinal data of specimen collection, as undertaken in a study on SARS-CoV-2 positivity over time^25^. Third, while our model accounted for time varying specimen collection, it did not account for the time interval between sample collection and laboratory testing. We assumed that all samples were tested immediately following collection, thereby neglecting the potential degradation of viral RNA and the consequent reduction in detectable viral loads over time. However, such degradation has recently been shown to be minimal^26^. Finally, the model focused on the general patients with LRTI and did not account for individual-level risk factors other than age in determining the RSV positive proportion in a given calendar month, such as comorbidities^27^.

## Conclusion and Public Health Implications

In summary, a novel Bayesian latent-class model was developed and validated to address the complex biases that arise in real-world RSV burden estimation using laboratory testing data. The model explicitly accounts for imperfect and varying sensitivity across diagnostic approaches, heterogeneous individual testing patterns, and temporal trends in viral detection. While conventional naïve and multiplier methods consistently underestimated RSV burden, the proposed Bayesian framework produced accurate and robust estimates when applied to sufficiently large datasets, specifically, when sample sizes reached 30,000 tests (corresponding to 15,000 patients) or more. Under such conditions, the model effectively disentangled true infection prevalence from measurement error, providing a more reliable basis for epidemiological inference. These findings establish a principled, flexible, and transparent analytical approach for integrating multi-source diagnostic data, setting a new standard for respiratory pathogen burden estimation in heterogeneous testing environments and supporting more rational vaccine strategies and public health decision-making.

## Ethics approval

Ethics approval is not required for statistical simulation studies.

## Author contributions

YL conceptualised the study, with contributions from HN and XW. BC led data collection, cleaning and analysis with contributions from DK, HZ and CW. YL, HN and XW provided critical oversight to data analysis and interpretation. BC wrote the first draft of the manuscript. All other authors contributed to data interpretation, and critically reviewed the manuscript. All authors read and approved the final manuscript and were responsible for the decision to submit the manuscript for publication.

## Conflicts of interest

YL is an employee of Nanjing Medical University, which was a paid consultant to Pfizer in connection with the conduct of the submitted work and development of this manuscript; YL reports grants from WHO, GSK, and MSD (all to institution), consulting fees from WHO, and support from Pfizer, MSD and Sanofi for attending meetings, outside the submitted work. HN is an employee of the University of Edinburgh, which received funding from Pfizer for the submitted work; HN reports grants from WHO, the Innovative Medicines Initiative, the National Institute for Health Research, MSD, Pfizer, and Icosavax /AstraZeneca; and personal fees from the Gates Foundation, Pfizer, GSK, Merck, Icosavax/AstraZeneca, and Sanofi outside the submitted work. EB, CL, AV and SU are Pfizer employees and may own Pfizer stock. All other authors declared that they have no competing interests.

## Disclosure Statement

The study data used was the outcome of collaboration between University of Bristol and Pfizer. University of Bristol was the study sponsor, and Pfizer is the funder.

## Data availability

The simulated data and code used in this analysis can be found in the following GitHub repository: https://github.com/cbb666/RSV_burden_bayesianStan.

## Use of artificial intelligence (AI) tools

No AI tools were used in preparing this manuscript.

## Reference

1. Cong B, Dighero I, Zhang T, Chung A, Nair H, Li Y. Understanding the age spectrum of respiratory syncytial virus associated hospitalisation and mortality burden based on statistical modelling methods: a systematic analysis. BMC Med. 2023 June 26;21(1):224.

2. Branche AR, Saiman L, Walsh EE, et al. Incidence of Respiratory Syncytial Virus Infection Among Hospitalized Adults, 2017-2020. Clin Infect Dis. 2022 Mar 23;74(6):1004–1011.

3. Rozenbaum MH, Begier E, Kurosky SK, et al. Incidence of respiratory syncytial virus infection in older adults: Limitations of current data. Infect Dis Ther. 2023 May 23;12(6):1487–1504.

4. CDC. RSV vaccine guidance for adults [Internet]. Respiratory Syncytial Virus Infection (RSV) 2025 [cited 2025 Dec 23]. Available from: https://www.cdc.gov/rsv/hcp/vaccine-clinical-guidance/adults.html

5. RSV vaccine [Internet]. nhs.uk 2024 [cited 2025 Dec 23]. Available from: https://www.nhs.uk/vaccinations/rsv-vaccine/

6. Recommendations by the standing committee on vaccination (STIKO) at the robert koch institute – 2025.

7. Rozenbaum MH, Judy J, Tran D, Yacisin K, Kurosky SK, Begier E. Low levels of RSV testing among adults hospitalized for lower respiratory tract infection in the united states. Infect Dis Ther. 2023 Jan 27;12(2):677–685.

8. Li K, Bont LJ, Weinberger DM, Pitzer VE. Relating in vivo respiratory syncytial virus infection kinetics to host infectiousness in different age groups. J Infect Dis. 2025 Sept 15;232(3):691–699.

9. Onwuchekwa C, Moreo LM, Menon S, et al. Underascertainment of Respiratory Syncytial Virus Infection in Adults Due to Diagnostic Testing Limitations: A Systematic Literature Review and Meta-analysis. J Infect Dis. 2023 Jan 20;228(2):173–184.

10. Onwuchekwa C, Atwell J, Moreo LM, et al. Pediatric Respiratory Syncytial Virus Diagnostic Testing Performance: A Systematic Review and Meta-analysis. J Infect Dis. 2023 June 7;228(11):1516–1527.

11. Walsh EE, Peterson DR, Kalkanoglu AE, Lee FE-H, Falsey AR. Viral shedding and immune responses to respiratory syncytial virus infection in older adults. J Infect Dis. 2013 May 1;207(9):1424–1432.

12. Begier E, Aliabadi N, Ramirez JA, et al. Detection by nasopharyngeal swabs alone underestimates respiratory syncytial virus–related hospitalization incidence in adults: The multispecimen study’s final analysis. J Infect Dis. 2025 Apr 18;232(1):e126–e136.

13. Ramirez J, Carrico R, Wilde A, et al. Diagnosis of respiratory syncytial virus in adults substantially increases when adding sputum, saliva, and serology testing to nasopharyngeal swab RT-PCR. Infect Dis Ther. 2023 May 6;12(6):1593–1603.

14. McLaughlin JM, Khan F, Begier E, Swerdlow DL, Jodar L, Falsey AR. Rates of Medically Attended RSV Among US Adults: A Systematic Review and Meta-analysis. Open Forum Infect Dis. 2022 June 17;9(7):ofac300.

15. Li Y, Kulkarni D, Begier E, et al. Adjusting for Case Under-Ascertainment in Estimating RSV Hospitalisation Burden of Older Adults in High-Income Countries: a Systematic Review and Modelling Study. Infect Dis Ther. 2023 Apr;12(4):1137–1149.

16. Havers FP, Whitaker M, Melgar M, et al. Burden of respiratory syncytial virus-associated hospitalizations in US adults, october 2016 to september 2023. JAMA Netw Open. 2024 Nov 13;7(11):e2444756.

17. Hak SF, Sankatsing VDV, Wildenbeest JG, et al. Burden of RSV infections among young children in primary care: A prospective cohort study in five european countries (2021-23). Lancet Respir Med. 2025 Feb;13(2):153–165.

18. Lihou K, Challen R, Chatzilena A, et al. Improving the accuracy of respiratory syncytial virus (RSV) incidence estimates among hospitalised adults in bristol, UK. BMC Infect Dis. 2025 Aug 21;25(1):1050.

19. UKHSA data dashboard [Internet]. [cited 2025 June 27]. Available from: https://ukhsa-dashboard.data.gov.uk

20. Hyams C, Begier E, Garcia Gonzalez M, et al. Incidence of acute lower respiratory tract disease hospitalisations, including pneumonia, among adults in bristol, UK, 2019, estimated using both a prospective and retrospective methodology. BMJ Open. 2022 June;12(6):e057464.

21. About the stan project [Internet]. Stan [cited 2025 Aug 14]. Available from: https://mc-stan.org/about/

22. Albert PS, Dodd LE. A cautionary note on the robustness of latent class models for estimating diagnostic error without a gold standard. Biometrics. 2004 June;60(2):427–435.

23. Smeden M van, Oberski DL, Reitsma JB, Vermunt JK, Moons KGM, Groot JAH de. Problems in detecting misfit of latent class models in diagnostic research without a gold standard were shown. J Clin Epidemiol. 2016 June;74:158–166.

24. Huggett JF, Benes V, Bustin SA, et al. Cautionary note on contamination of reagents used for molecular detection of SARS-CoV-2. Clin Chem. 2020 Oct 30;66(11):1369–1372.

25. Overton CE, Fyles M, Mellor J, et al. SARS-CoV-2 test sensitivity and duration of positivity in the UK during the 2023/2024 winter: A prospective cohort study based on self-reported data. J Infect. 2025 June 1;90(6):106485.

26. Lihou K, McGuinness S, Morales-Aza B, et al. Sensitivity of RSV detection by PCR in respiratory samples is not reduced by a 24[2h delay from sampling to testing with storage at room temperature. Mol Med. 2025 Nov 22;31(1):342.

27. Shi T, Vennard S, Jasiewicz F, Brogden R, Nair H, RESCEU Investigators. Disease Burden Estimates of Respiratory Syncytial Virus related Acute Respiratory Infections in Adults With Comorbidity: A Systematic Review and Meta-Analysis. The Journal of Infectious Diseases. 2022 Aug 1;226(Supplement_1):S17–S21.

28. World Population Prospects 2022: Summary of Results | Population Division [Internet]. [cited 2024 Feb 18]. Available from: https://www.un.org/development/desa/pd/content/World-Population-Prospects-2022

29. COVID-19 & respiratory surveillance [Internet]. [cited 2025 June 27]. Available from: https://scotland.shinyapps.io/phs-respiratory-covid-19/

