## Supplementary file for "A Bayesian latent-class model framework to estimate disease burden of respiratory syncytial virus using imperfect and heterogeneous laboratory diagnostic data"

### Supplementary methods

#### Text S1. Generation of the true values of age- and calendar-month specific RSV positive proportion.

As data on age- and calendar-month specific RSV positive proportion in UK were not available, we calculated the calibrated RSV positive proportion as an alternative for the corresponding true values, as follows –

$${Prop}_{m,a}=\frac{{MeanProp}_{a}}{{MeanRate}_{a}} \times{Rate}_{m,a} (4)$$

${Prop}_{m,a}$ represented the calibrated RSV positive proportion among LRTI cases in a given age group $a$ and month $m$ during the study period (i.e., August 1, 2021 to July 2024). $Mean{Prop}_{a}$ was age-specific RSV positive proportion among LRTI patients for 2022–2023 season in Bristol, UK (available from a published study)^1^. ${Rate}_{m,a}$ and ${MeanRate}_{a}$ were age- and calendar-month specific RSV incidence rate and the average age-specific RSV incidence rate in the community during the study period, respectively (available from the Scottish surveillance dashboard)^2^.

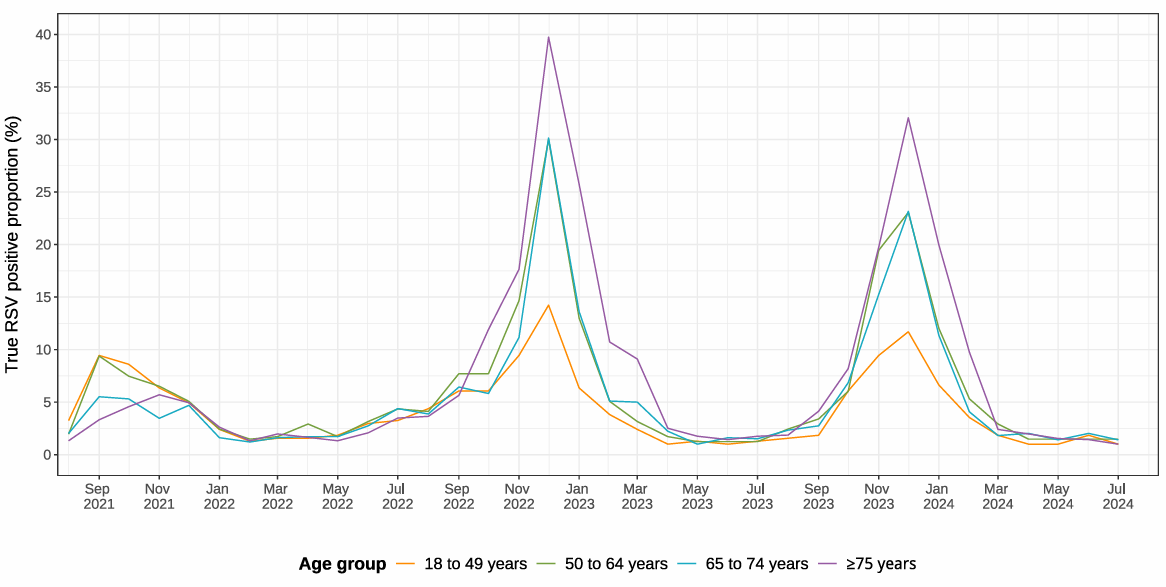

#### Figure S1. The pre-specified true values of age- and calendar-month specific RSV positive proportion.

RSV: respiratory syncytial virus; LRTI: lower respiratory tract infection. The calculation process was shown in **Text S1**.

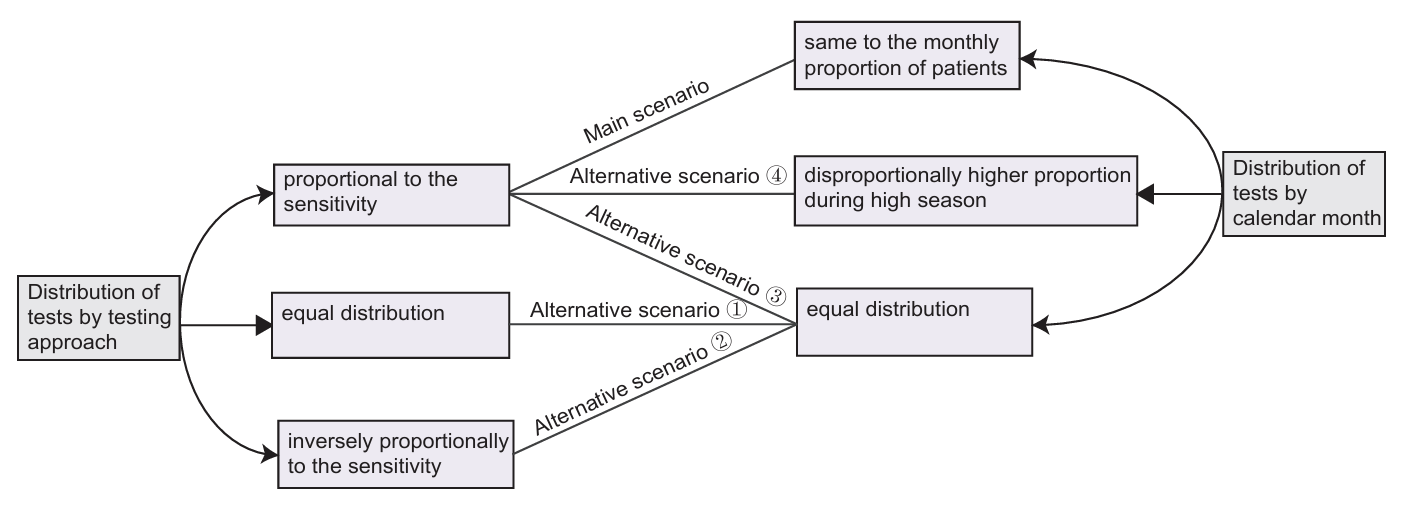

#### Figure S2. Combinations of distribution of tests by testing approach and calendar month for generating simulated datasets.

#### Table S1. Practical sensitivity estimates of different combinations of testing approaches by the multiplier approach under varied sample size scenarios, presented as range across four simulated datasets.

| **Practical sensitivity (%) by combination of testing approaches (Test 1 – Test 4)** | | | | | | | | | | | | | | | |
| --- | --- | --- | --- | --- | --- | --- | --- | --- | --- | --- | --- | --- | --- | --- | --- |
| **Test 1** | ⬤ | ⬤ |  |  |  | ⬤ |  |  | ⬤ | ⬤ |  | ⬤ |  | ⬤ | ⬤ |
| **Test 2** | ⬤ |  | ⬤ |  |  | ⬤ | ⬤ |  |  |  | ⬤ | ⬤ | ⬤ |  | ⬤ |
| **Test 3** | ⬤ |  |  | ⬤ |  |  | ⬤ | ⬤ |  | ⬤ |  | ⬤ | ⬤ | ⬤ |  |
| **Test 4** | ⬤ |  |  |  | ⬤ |  |  | ⬤ | ⬤ |  | ⬤ |  | ⬤ | ⬤ | ⬤ |
| **Expected value*** | 100 | 81 | 71 | 61 | 51 | 95 | 89 | 81 | 91 | 93 | 86 | 99 | 95 | 97 | 98 |
| **Estimates under varied sample size scenarios** | | | | | | | | | | | | | | | |
| **2,500** | ref | 66–81 | 63–71 | 51–61 | 46–51 | 81–87 | 77–84 | 63–72 | 73–80 | 76–83 | 71–77 | 94–97 | 85–90 | 84–88 | 87–94 |
| **5,000** | ref | 70–74 | 62–71 | 54–61 | 47–56 | 78–87 | 76–78 | 68–75 | 76–80 | 78–81 | 73–78 | 90–94 | 86–91 | 86–91 | 90–93 |
| **7,500** | ref | 74–77 | 62–68 | 57–61 | 49–58 | 80–87 | 76–79 | 69–75 | 77–83 | 81–86 | 70–77 | 92–94 | 84–88 | 90–92 | 87–92 |
| **10,000** | ref | 70–77 | 63–69 | 52–63 | 50–54 | 82–85 | 75–79 | 66–75 | 74–80 | 80–82 | 73–76 | 92–94 | 84–89 | 88–92 | 89–91 |
| **15,000** | ref | 71–77 | 64–70 | 55–62 | 50–57 | 83–87 | 76–78 | 69–72 | 79–79 | 79–85 | 75–79 | 92–93 | 86–87 | 87–91 | 90–94 |
| **20,000** | ref | 70–76 | 63–68 | 54–59 | 47–54 | 82–86 | 75–78 | 68–71 | 77–82 | 79–82 | 74–76 | 91–93 | 84–88 | 87–90 | 91–92 |
| **30,000** | ref | 72–74 | 66–67 | 58–61 | 50–52 | 85–85 | 77–80 | 70–71 | 77–79 | 81–82 | 74–76 | 93–94 | 86–87 | 87–90 | 91–93 |

*Derived based on conditional probability formula: $P\left( A | B \right)=\frac{P(A\cap B)}{P(B)}$, where B denotes the gold standard defined by the multiplier approach and A denotes a specific combination of testing approach. Note that the gold standard is a practical definition and does not include RSV infected cases that are missed by all of the four testing approaches; therefore, the sensitivity estimates as defined in the multiplier approach differs from the true sensitivity, i.e., $P\left( A | B \right)\neq P(A)$.

### Supplementary results

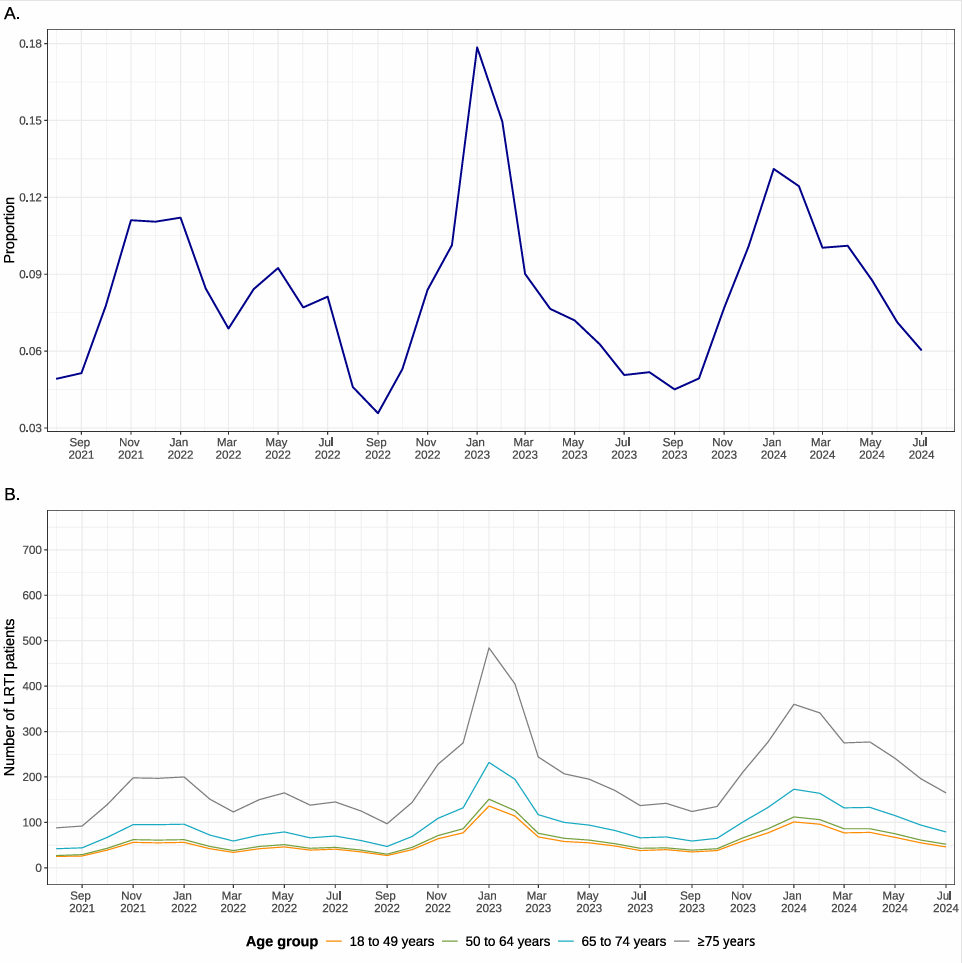

#### Figure S3. The distribution of LRTI patients by calendar month in the UK from UKSHA dashboard (A) and by age group and calendar month for simulated dataset (B).

LRTI: lower respiratory tract infection. The sample size scenario of 15,000 LRTI patients and 30,000 tests was used for the simulated dataset. Note that the values on the Y-axis represents the relative proportion of incidence rate of LRTI for each calendar month (from August 1, 2021 to July 31, 2024 in the UK); therefore, the sum of the relative proportions of the 36 months is 1.

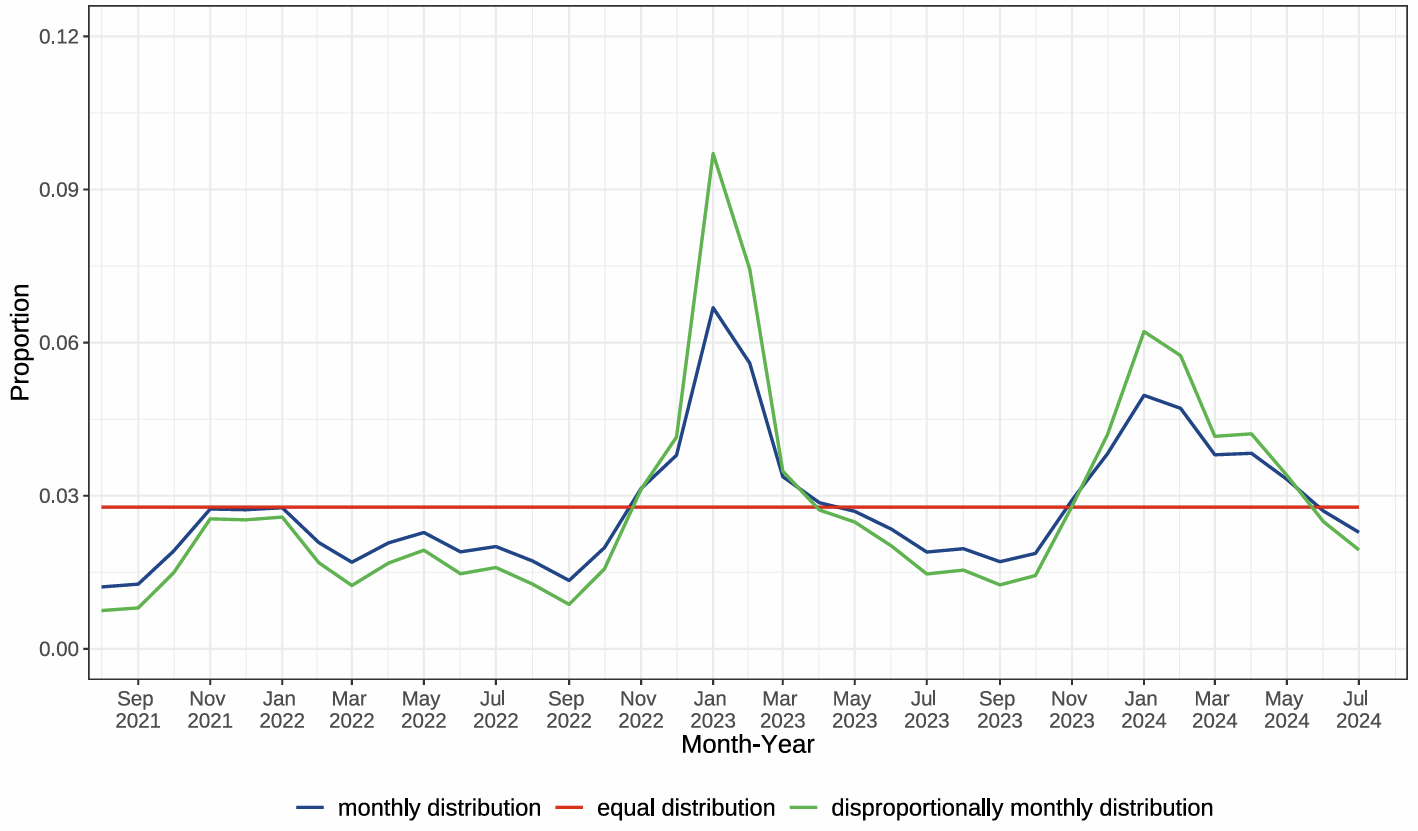

#### Figure S4. Distribution of tests by calendar month for simulated dataset by scenario.

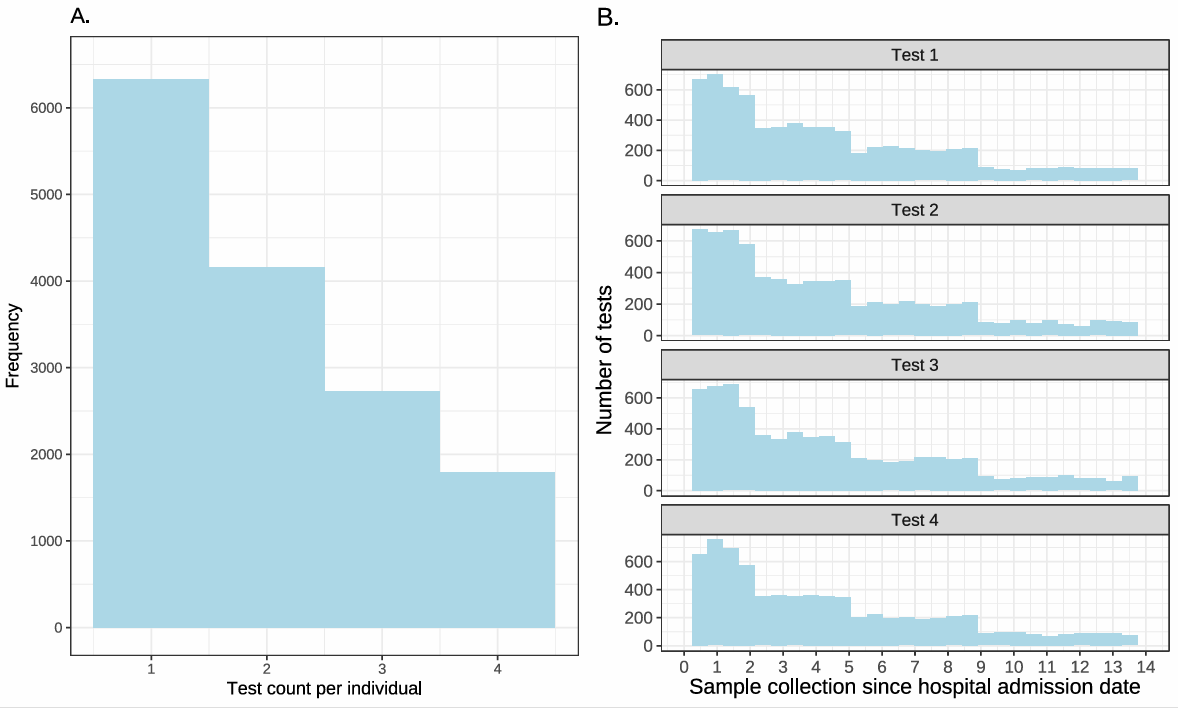

#### Figure S5. Distribution of number of tests per patient (A) and specimen collection date since hospital admission date (B) for simulated dataset.

The sample size scenario of 15,000 LRTI patients and 30,000 tests was used for the simulated dataset.

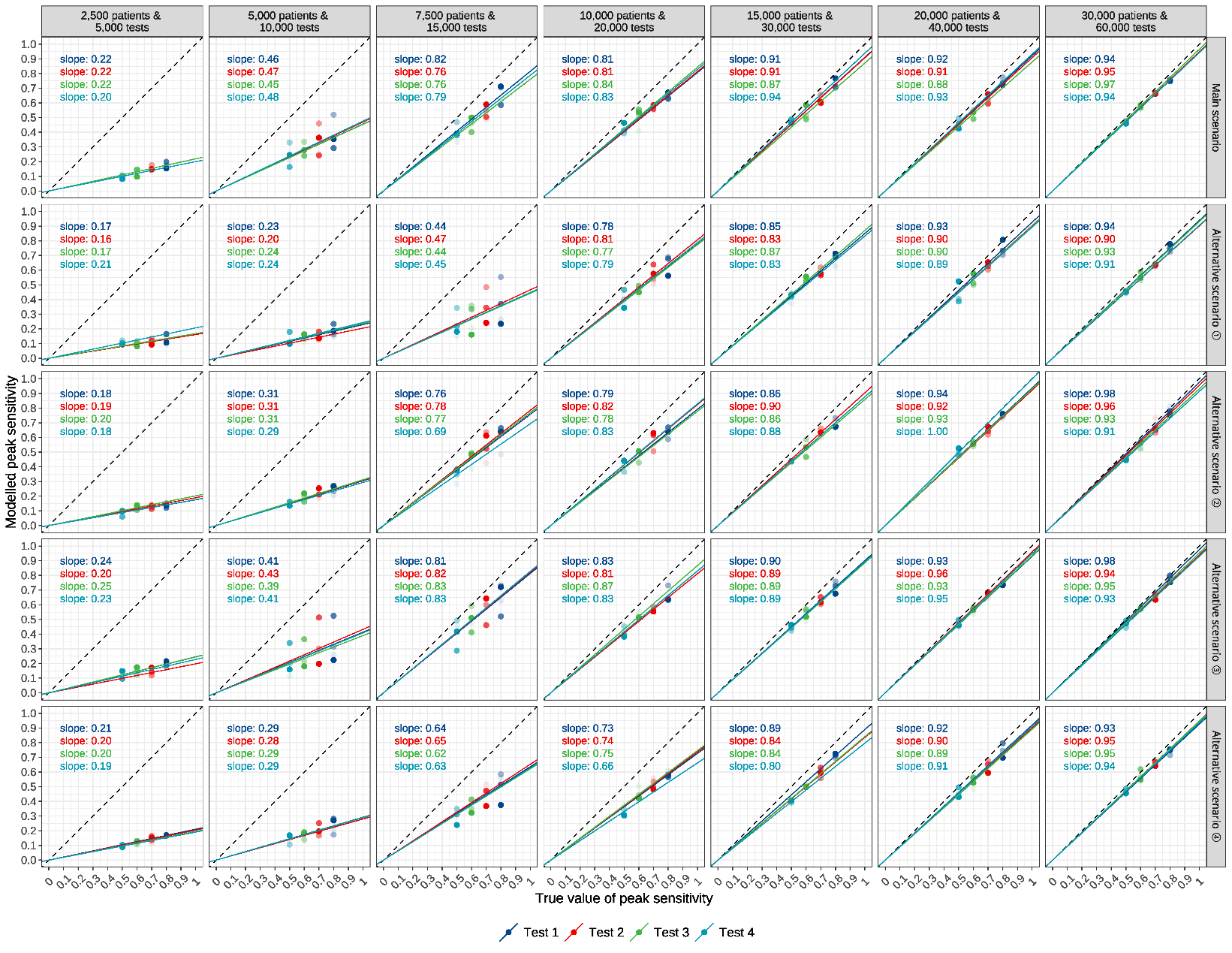

#### Figure S6. Comparison of model estimates and true values of peak sensitivity of tests, under varied sample size and testing practice scenarios.

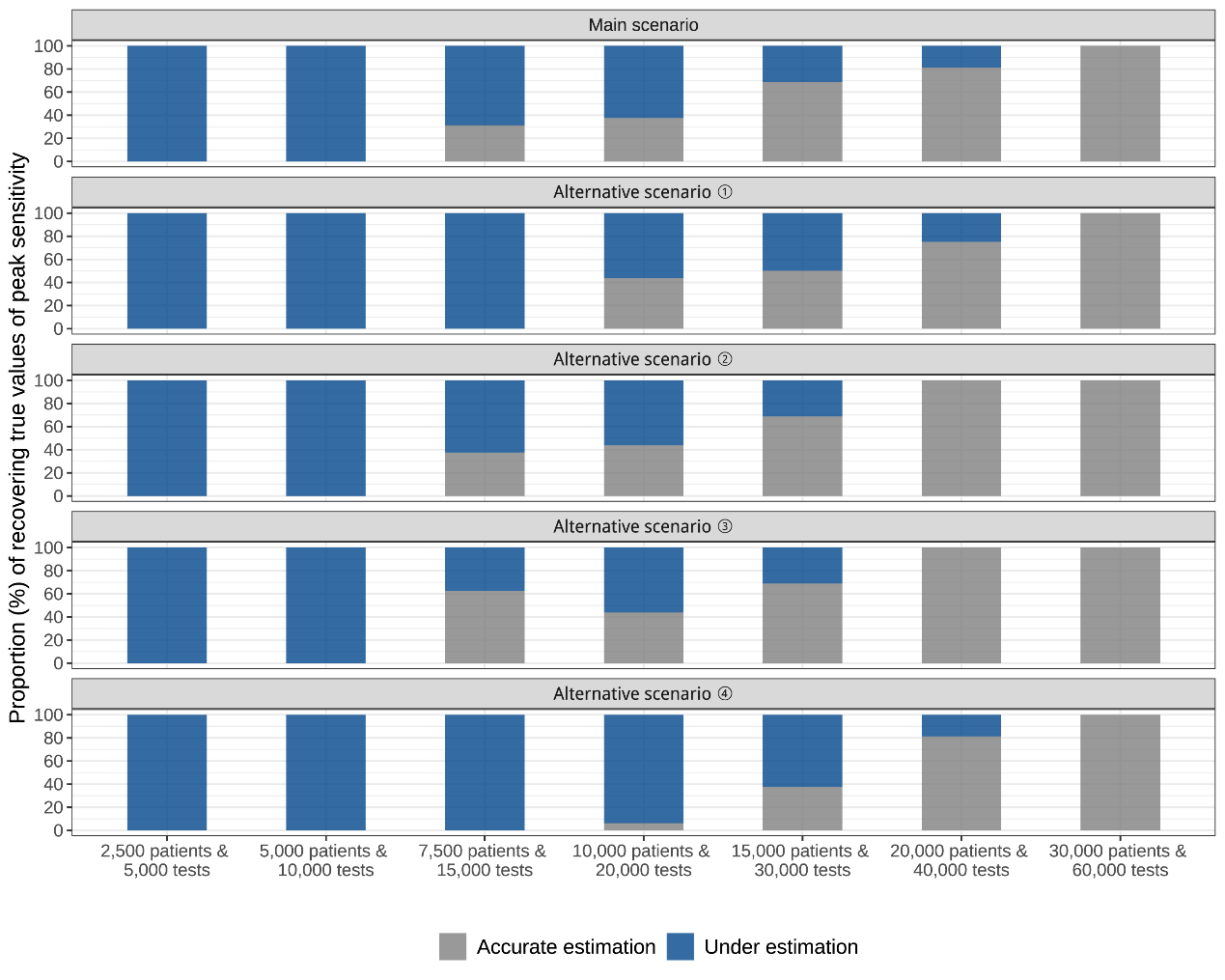

#### Figure S7. Proportion of estimated 95% credible intervals for model parameters containing true values of peak sensitivity of tests, under varied sample size and testing practice scenarios.

Note that four simulated datasets were generated for each combination of sample size and testing practice scenario. Accurate estimation is defined as the 95% credible intervals containing true values; under [over] estimation denotes the upper [lower] limit of the 95% credible intervals was lower [higher] than the true value.

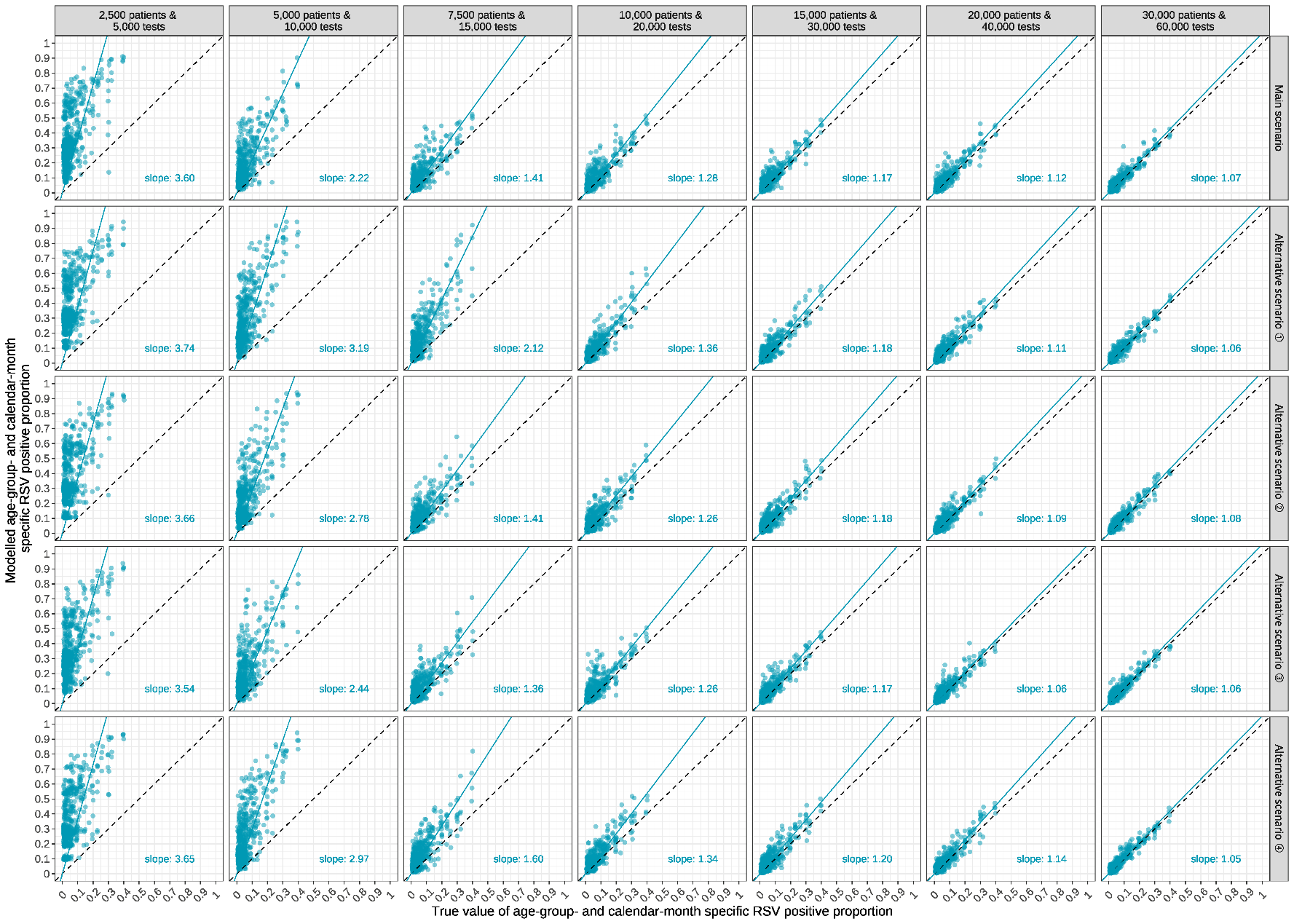

#### Figure S8. Comparison of model estimates and true values of age- and calendar-month specific RSV positive proportion, under varied sample size and testing practice scenarios.

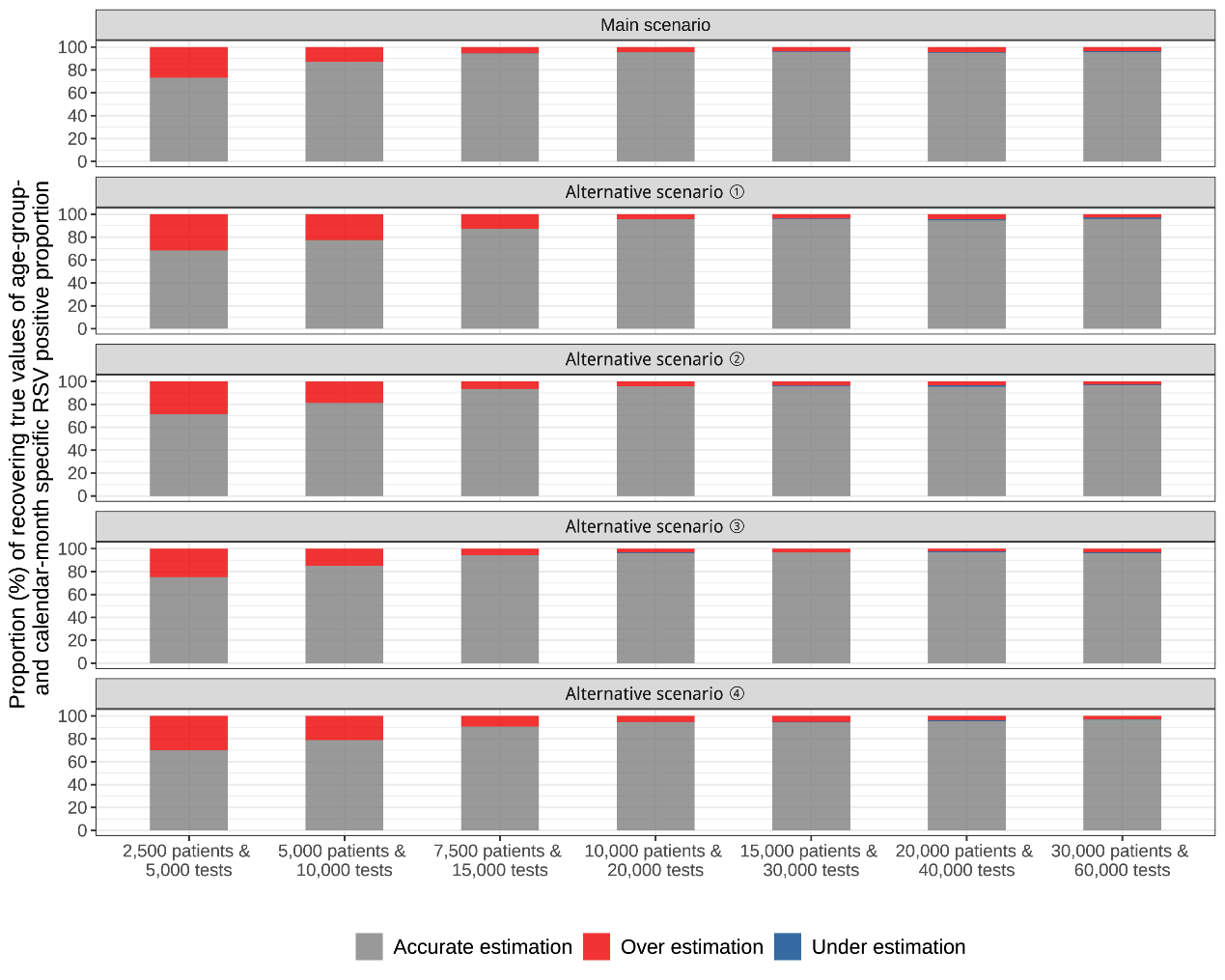

#### Figure S9. Proportion of estimated 95% credible intervals for model parameters containing true values of age- and calendar-month specific RSV positive proportion, under varied sample size and testing practice scenarios.

Accurate estimation is defined as the 95% credible intervals containing true values; under [over] estimation denotes the upper [lower] limit of the 95% credible intervals was lower [higher] than the true value.

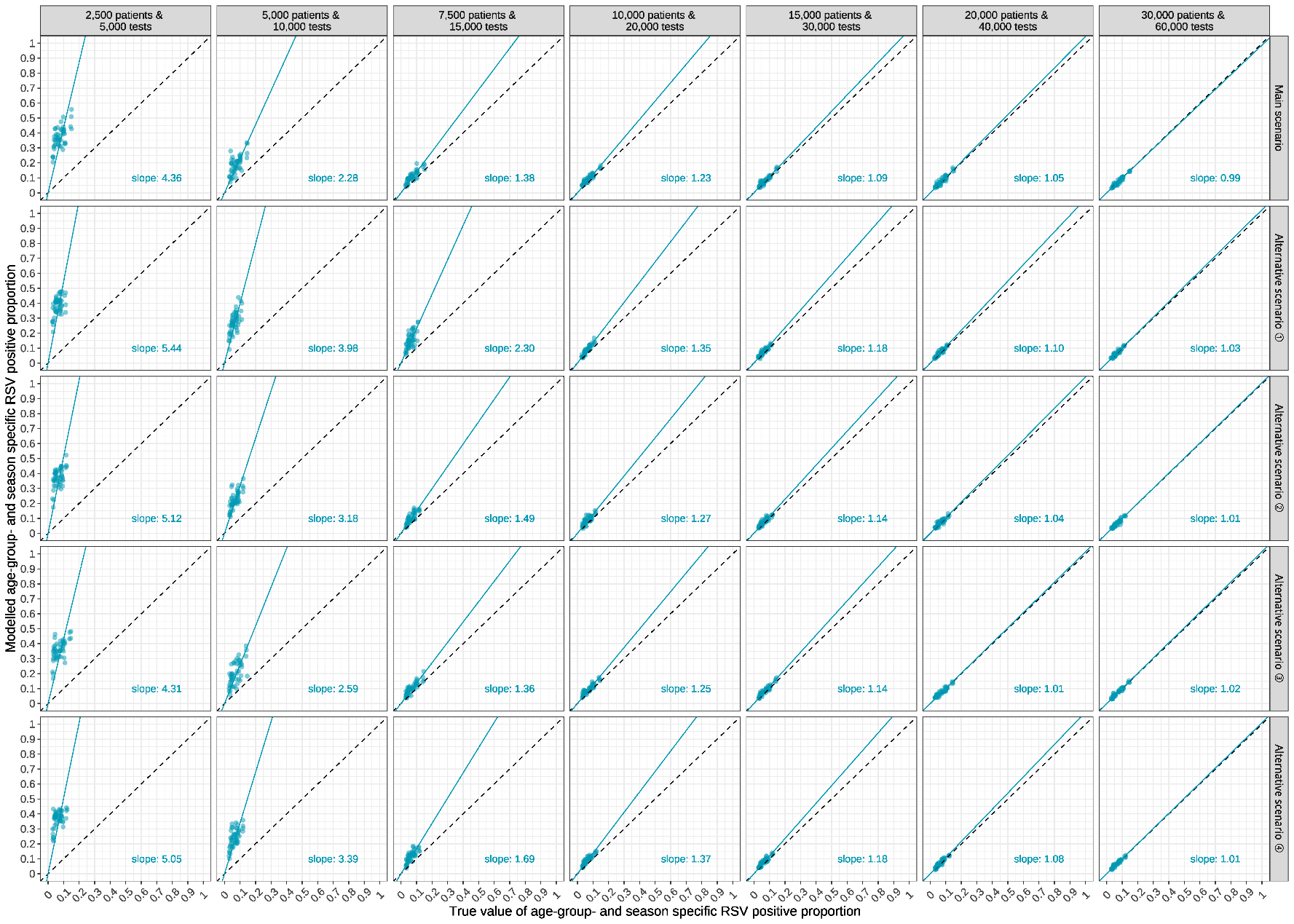

#### Figure S10. Comparison of model estimates and true values of age- and season specific RSV positive proportion, under varied sample size and testing practice scenarios.

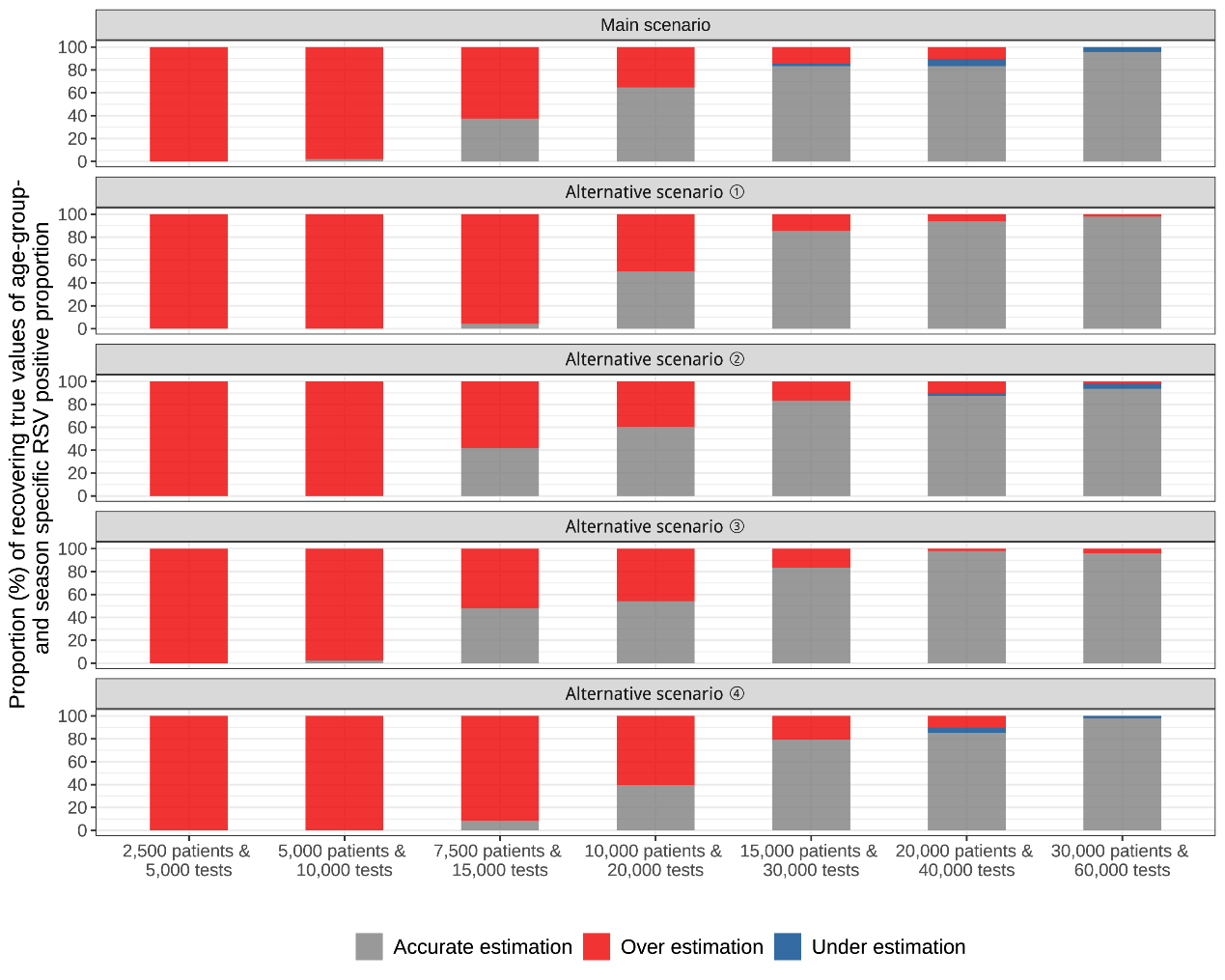

#### Figure S11. Proportion of estimated 95% credible intervals for model parameters containing true values of age- and season-specific RSV positive proportion, under varied sample size and testing practice scenarios.

Accurate estimation is defined as the 95% credible intervals containing true values; under [over] estimation denotes the upper [lower] limit of the 95% credible intervals was lower [higher] than the true value.

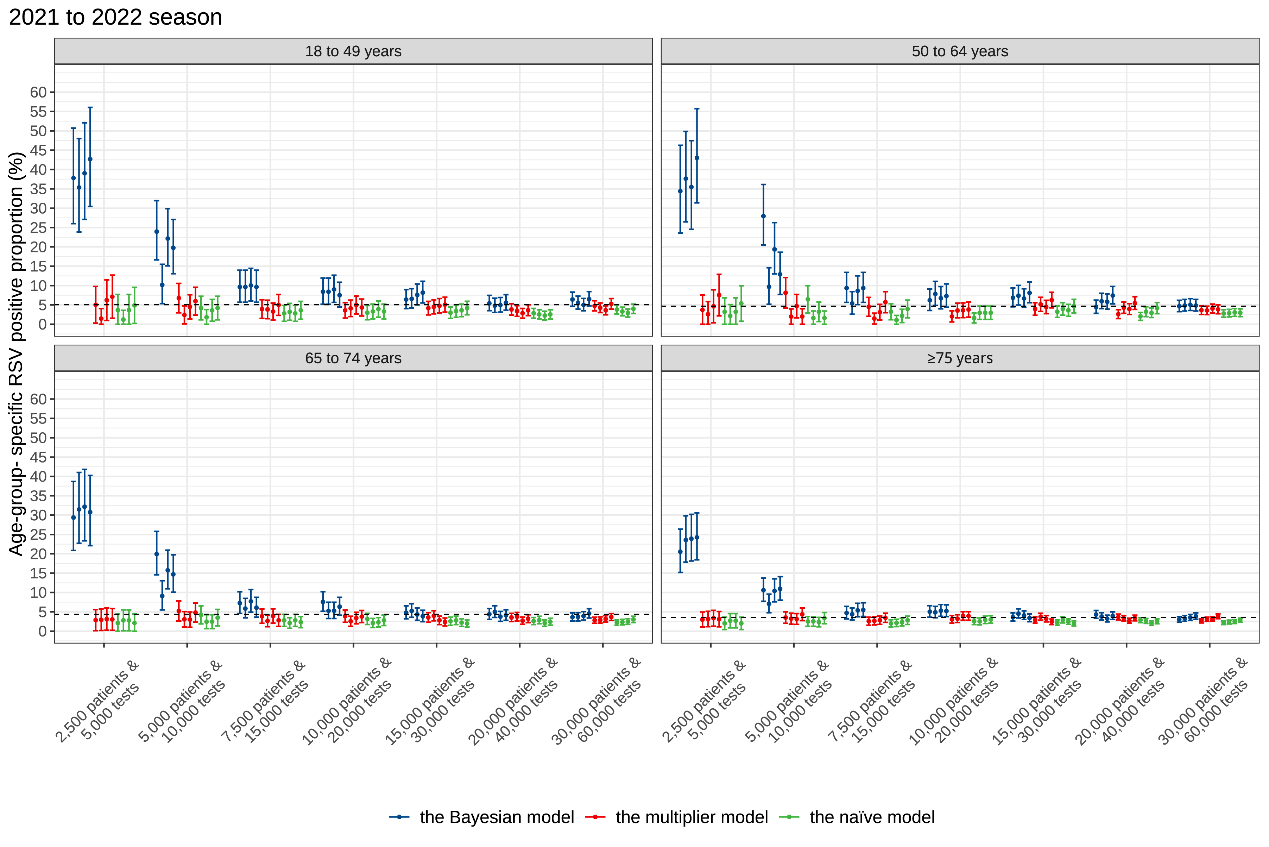

#### Figure S12. Comparison of age-specific RSV positive proportion from August 1, 2021 to July 31, 2022 among the Bayesian model, naïve model, and multiplier model under varied sample size scenarios.

Four simulated datasets were generated for each sample size scenario. Dotted lines represent true values of RSV positive proportions. Dots indicate modelled point estimates and corresponding error bars indicate corresponding 95% credible intervals or confidence intervals.

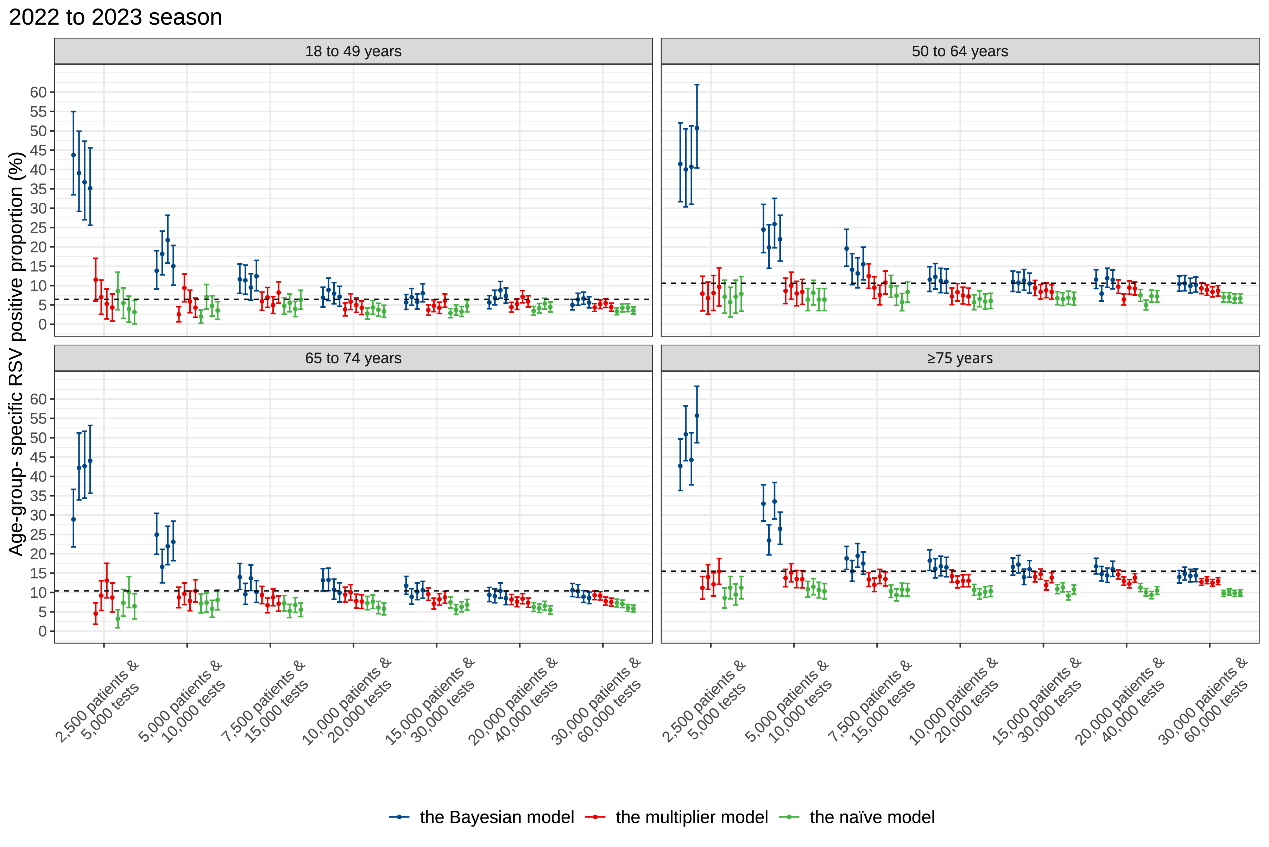

#### Figure S13. Comparison of age-specific RSV positive proportion from August 1, 2022 to July 31, 2023 among the Bayesian model, naïve model, and multiplier model under varied sample size scenarios.

Four simulated datasets were generated for each sample size scenario. Dotted lines represent true values of RSV positive proportions. Dots indicate modelled point estimates and corresponding error bars indicate corresponding 95% credible intervals or confidence intervals.

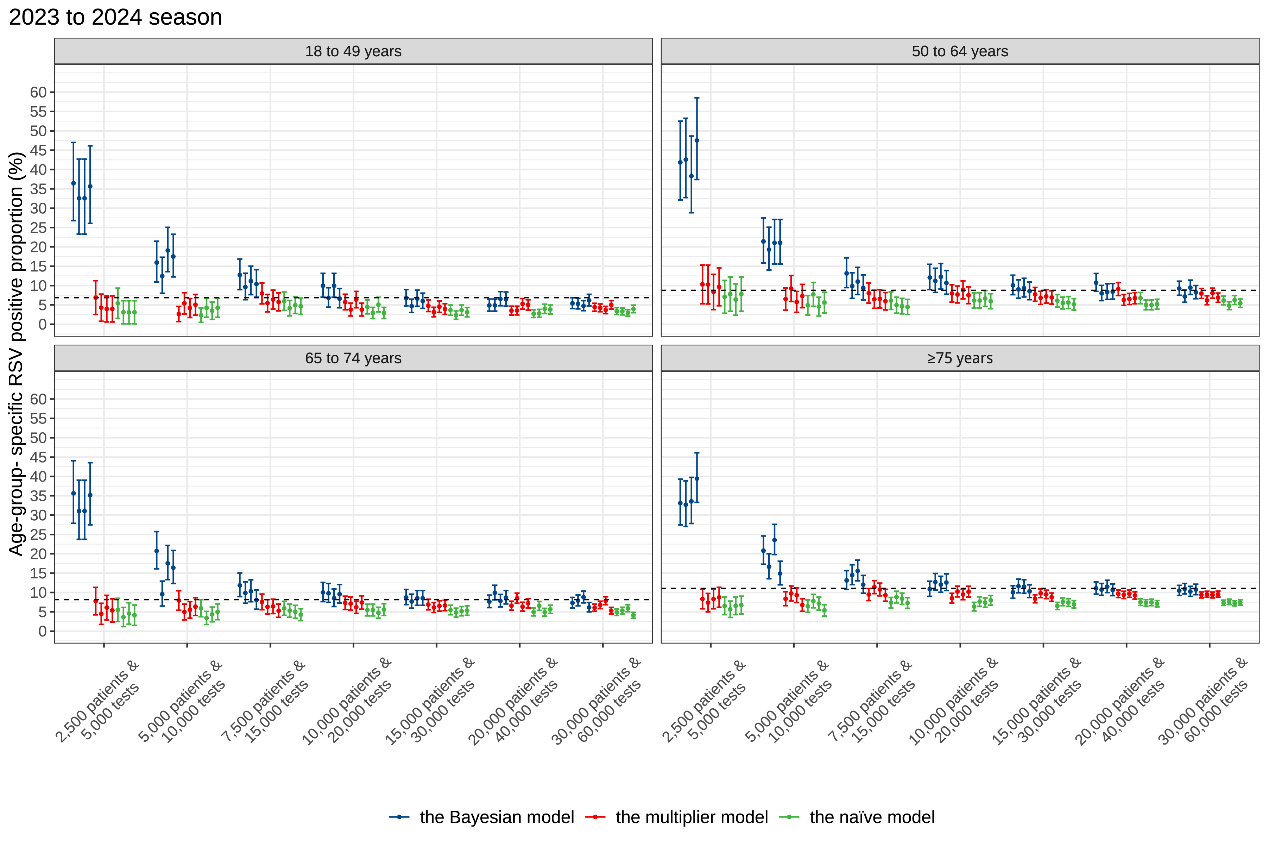

#### Figure S14. Comparison of age-specific RSV positive proportion from August 1, 2023 to July 31, 2024 among the Bayesian model, naïve model, and multiplier model under varied sample size scenarios.

Four simulated datasets were generated for each sample size scenario. Dotted lines represent true values of RSV positive proportions. Dots indicate modelled point estimates and corresponding error bars indicate corresponding 95% credible intervals or confidence intervals.

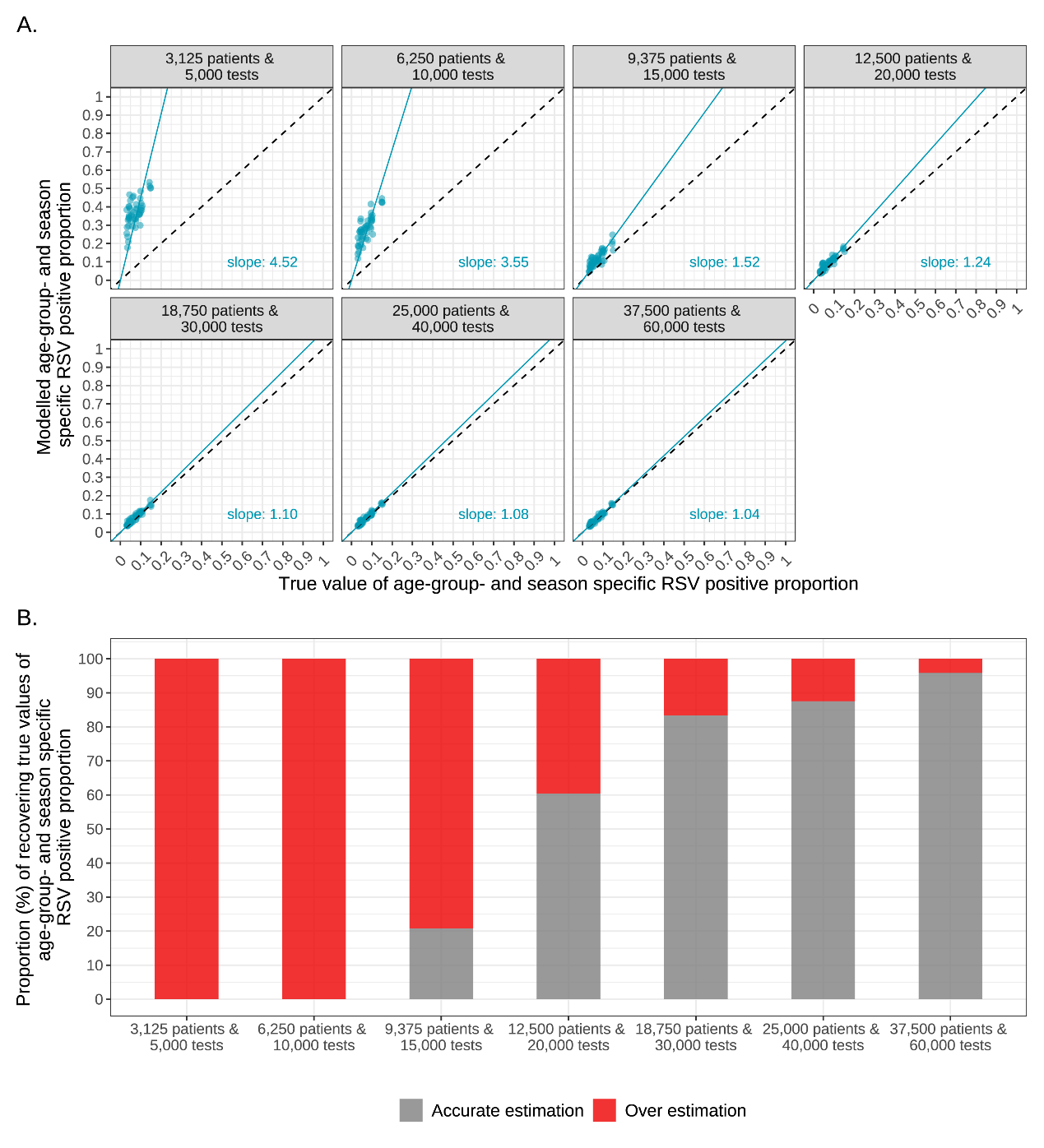

#### Figure S15. Comparison of model estimates and true values of age- and season- specific RSV positive proportion under varied sample size scenarios, in an exploratory analysis that retained 3 testing approaches.

Four simulated datasets were generated for each sample size scenario. Accurate estimation is defined as the 95% credible intervals containing the true values; under [over] estimation denotes the upper [lower] limit of the 95% credible intervals being lower [higher] than the true value.

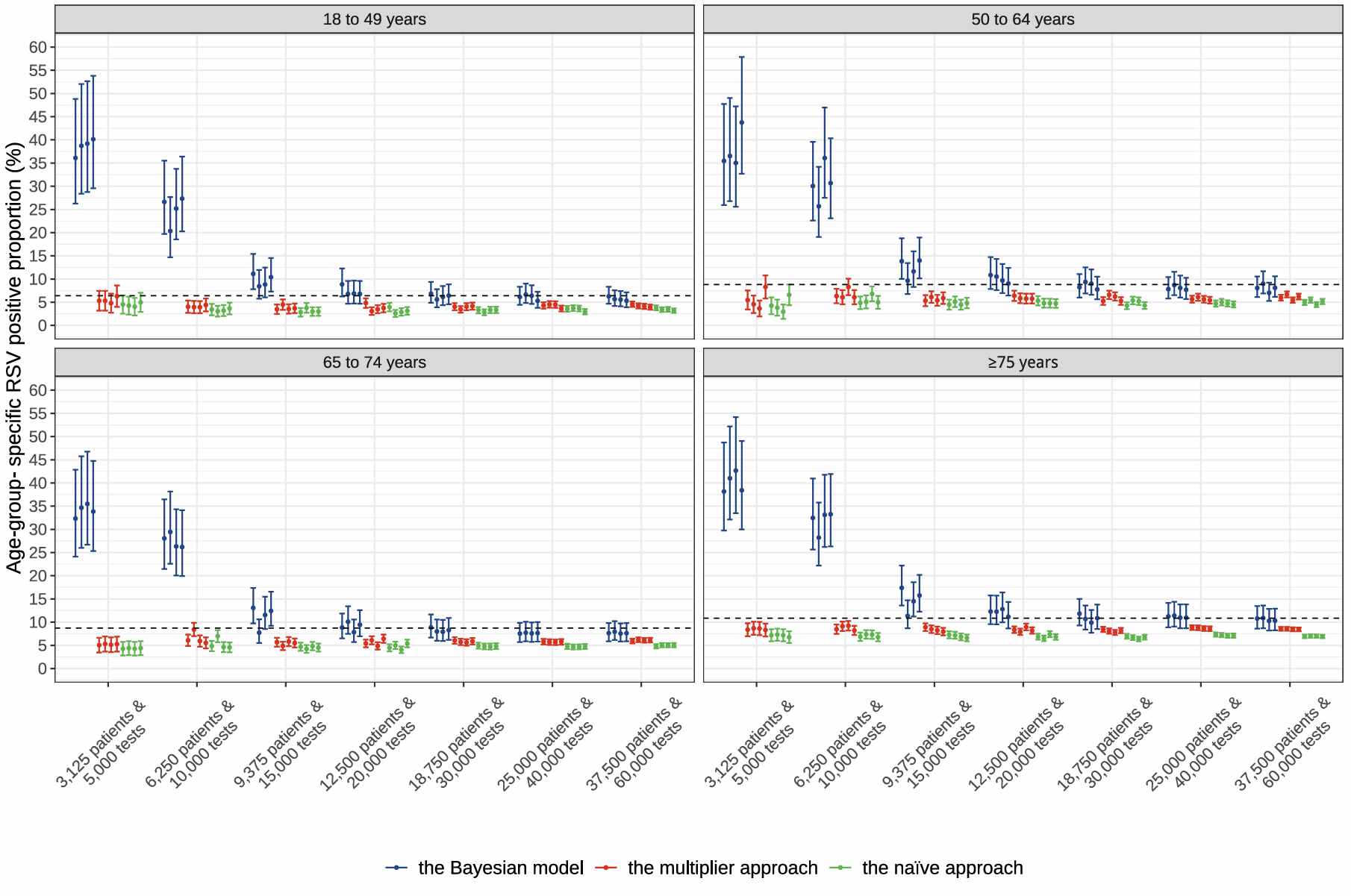

#### Figure S16. Comparison of age- specific annual RSV positive proportion from August 1, 2021 to July 31, 2024 among the Bayesian model, naïve model, and multiplier model under varied sample size scenarios, in an exploratory analysis that retained 3 testing approaches.

Four simulated datasets were generated for each sample size scenario. Dotted lines represent true values of RSV positive proportions. Dots indicate modelled point estimates and corresponding error bars indicate corresponding 95% credible intervals or confidence intervals.

### Reference

1.	Lihou K, Challen R, Chatzilena A, et al. Improving the accuracy of respiratory syncytial virus (RSV) incidence estimates among hospitalised adults in bristol, UK. *BMC Infect Dis*. 2025 Aug 21;**25**(1):1050.

2.	COVID-19 & respiratory surveillance [Internet]. [cited 2025 June 27]. Available from: https://scotland.shinyapps.io/phs-respiratory-covid-19/
